# The effect of transcutaneous auricular vagus nerve stimulation on cardiovascular function in subarachnoid hemorrhage patients: a safety study

**DOI:** 10.1101/2024.04.03.24304759

**Authors:** Gansheng Tan, Anna L. Huguenard, Kara M. Donovan, Phillip Demarest, Xiaoxuan Liu, Ziwei Li, Markus Adamek, Kory Lavine, Ananth K. Vellimana, Terrance T. Kummer, Joshua W. Osbun, Gregory J. Zipfel, Peter Brunner, Eric C. Leuthardt

## Abstract

**Introduction:** Subarachnoid hemorrhage (SAH) is characterized by intense central inflammation, leading to substantial post-hemorrhagic complications such as vasospasm and delayed cerebral ischemia. Given the anti-inflammatory effect of transcutaneous auricular vagus nerve stimulation (taVNS) and its ability to promote brain plasticity, taVNS has emerged as a promising therapeutic option for SAH patients. However, the effects of taVNS on cardiovascular dynamics in critically ill patients, like those with SAH, have not yet been investigated. Given the association between cardiac complications and elevated risk of poor clinical outcomes after SAH, it is essential to characterize the cardiovascular effects of taVNS to ensure this approach is safe in this fragile population. Therefore, we assessed the impact of both acute taVNS and repetitive taVNS on cardiovascular function in this study.

**Methods:** In this randomized clinical trial, 24 SAH patients were assigned to either a taVNS treatment or a Sham treatment group. During their stay in the intensive care unit, we monitored patient electrocardiogram (ECG) readings and vital signs. We compared long-term changes in heart rate, heart rate variability, QT interval, and blood pressure between the two groups. Additionally, we assessed the effects of acute taVNS by comparing cardiovascular metrics before, during, and after the intervention. We also explored acute cardiovascular biomarkers in patients exhibiting clinical improvement.

**Results:** We found that repetitive taVNS did not significantly alter heart rate, QT interval, blood pressure, or intracranial pressure. However, taVNS increased overall heart rate variability and parasympathetic activity compared to the sham treatment. The increase in parasympathetic activity was most pronounced from 2–4 days after initial treatment (Cohen’s d = 0.50). Acutely, taVNS increased heart rate, blood pressure, and peripheral perfusion index without affecting the corrected QT interval, intracranial pressure, or heart rate variability. The acute post-treatment elevation in heart rate was more pronounced in patients who experienced a decrease of more than one point in their Modified Rankin Score at the time of discharge.

**Conclusions:** Our study found that taVNS treatment did not induce adverse cardiovascular effects, such as bradycardia or QT prolongation, supporting its development as a safe immunomodulatory treatment approach for SAH patients. The observed acute increase in heart rate after taVNS treatment may serve as a biomarker for SAH patients who could derive greater benefit from this treatment.

**Trial registration:** NCT04557618

## 1. Introduction

Subarachnoid hemorrhage (SAH) is a devastating subtype of stroke that represents a significant global health burden and causes permanent disability in approximately 30% of survivors.^1,6,45^ Early brain injury can occur within the first 24 to 48 hours after ictus, which involves a cascade of elevated intracranial pressure and a subsequent drop of cerebral perfusion.^4^ Systemic and local inflammation, cerebral edema, blood-brain barrier (BBB) disruption, sympathetic nervous system activation, autoregulatory failure, microthrombosis, spreading depolarizations (SDs), and inflammation have all been observed during this period.^11,12^ These biological processes result in the inability of cerebral perfusion to match metabolic demands, leading to secondary brain injury and delayed cerebral ischemia that typically occurs between 5 and 14 days after the SAH.^2,7,8,9^ Delayed cerebral ischemia and deleterious inflammation are major predictors of poor outcomes and morbidity. The autonomic nervous system (ANS), comprising the sympathetic and the parasympathetic nervous system, plays a critical role in maintaining physiological homeostasis. SAH is believed to cause sympathetic predominance, which plays a key role in the development of cerebral vasospasm, renders patients more susceptible to non-neurological complications, and exacerbates deleterious inflammatory processes.

Numerous interventions have been explored to address the complex pathologies of subarachnoid hemorrhage (SAH) that contribute to secondary brain injury, aiming to improve patient outcomes.^14^ Transcutaneous auricular vagus nerve stimulation (taVNS) is one of the most promising therapeutic options, as recent studies have demonstrated its efficacy in reducing inflammation, improving autonomic balance, and enhancing brain plasticity.^3,10,13,14^ The auricular branch of the vagus nerve is a sensory nerve that innervates the external ear, including the cymba concha. Stimulating the auricular branch of the vagus nerve has been shown to activate the same brain regions as cervical vagus nerve stimulation^16^. Specifically, taVNS mediates cholinergic signaling and regulates proinflammatory responses via the inflammatory reflex (**Figure 1A**).^17,18^ In this reflex, inflammatory mediators such as cytokines trigger afferent vagus nerve signaling. This afferent signal then prompts an efferent response from the vagus nerve that acts to reduce the production of pro-inflammatory cytokines.^19^

**Figure 1.**
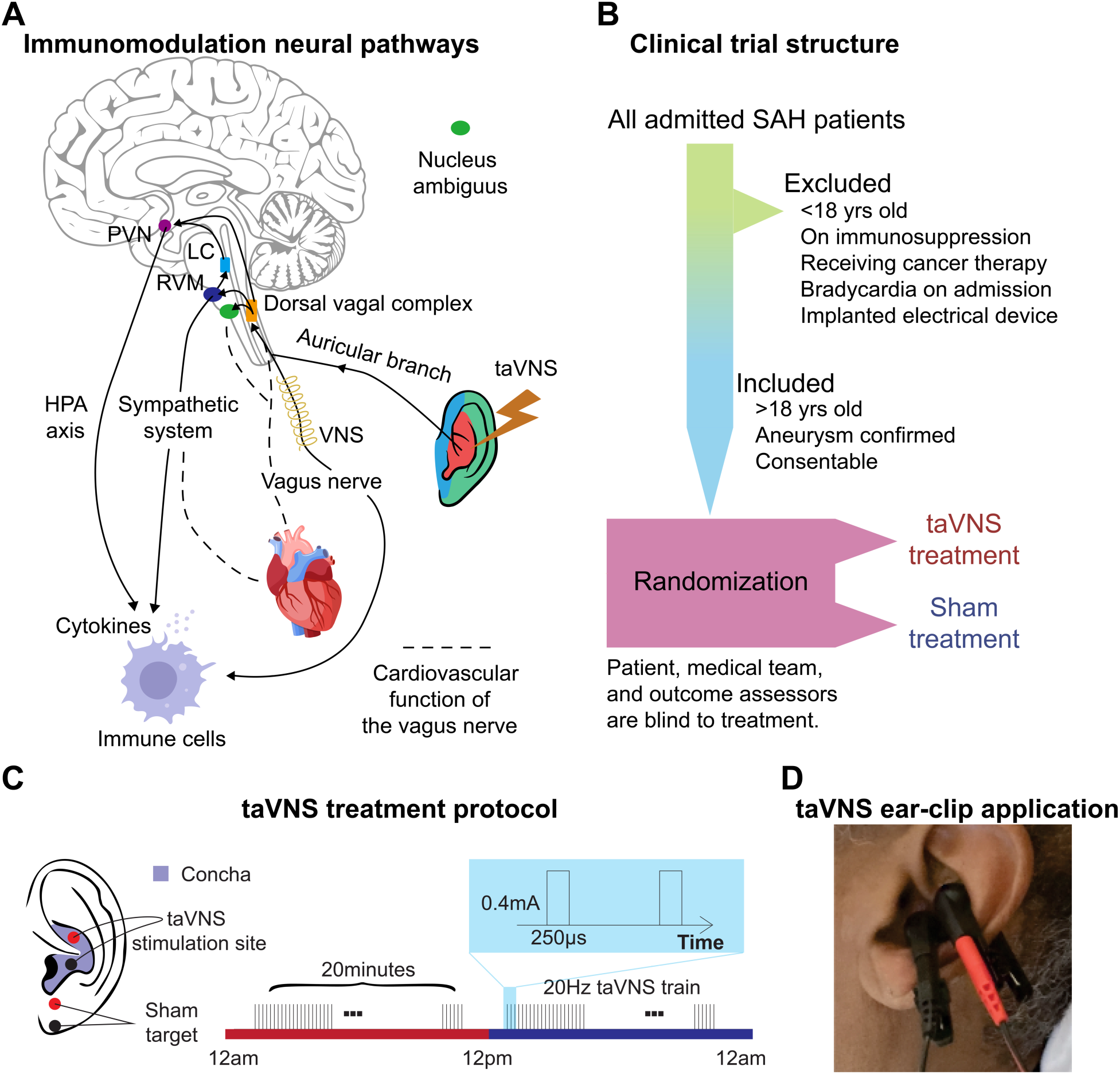
Study rationale and clinical trial design. **A.** Immunomodulation neural pathways associated with vagus nerve stimulation include cholinergic anti-inflammatory pathway, sympathetic nervous system, and hypothalamic-pituitary-adrenal (HPA) axis. Immunogenic stimuli activate vagal afferents terminating primarily in the dorsal vagal complex. Ascending projections from the dorsal vagal complex reach the paraventricular nucleus (PVN) and rostral ventromedial medulla (RVM), activating the hypothalamic-pituitary-adrenal (HPA) axis and sympathetic system, respectively, to regulate the immune response. taVNS can affect cardiovascular function through the sympathetic system or efferent vagus nerve. **B**-**C**. Clinical trial structure and treatment protocol. Patients in the taVNS group received electrical stimulation (0.4 mA, 250 µs pulse width, 20 Hz) for 20 minutes twice daily. Sham group patients wore the ear clip on the earlobe for the same duration. **D.** Ear clip application for taVNS treatment.

The vagus nerve also mediates cardiovascular function by regulating the autonomic system and metabolic homeostasis (**Figure 1A**).^20^ Theoretically, taVNS increases parasympathetic activity, which can be measured as increased heart rate variability (HRV). While some animal studies have reported a potential risk of bradycardia and decreased blood pressure associated with vagus nerve stimulation, two reviews of human studies have considered the cardiovascular effects of taVNS generally safe, with adverse effects reported only in patients with pre-existing heart diseases. ^21,22,23^ However, its cardiovascular effect in SAH patients is largely unknown. Given that critically ill patients, such as those with SAH, are extremely vulnerable to cardiovascular complications, it is essential to thoroughly examine the cardiovascular implications of taVNS to ensure its safety in this fragile population. This is particularly notable as cardiovascular abnormalities following SAH, such as prolonged elevated heart rate and QT prolongation, are associated with an increased risk of poor outcomes^5,24,25^. However, our limited understanding of these effects constitutes a significant barrier, preventing the advancement of taVNS from a promising therapeutic approach to an established clinical treatment for SAH. To address this gap, we assessed the effects of acute and repetitive taVNS on cardiovascular function based on electrocardiogram (ECG) and other monitored vital signs from SAH patients in the intensive care unit (ICU). The current study is part of the NAVSaH trial (NCT04557618) and focuses on the trial’s secondary outcomes, including heart rate, QT interval, HRV, and blood pressure.^32^ This interim analysis aims to evaluate the cardiovascular safety of the taVNS protocol and to provide insights that will inform the application of taVNS in SAH patients. The primary outcomes of this trial, including change in the inflammatory cytokine TNF-α and rate of radiographic vasospasm, are available as a pre-print and currently under review.^26^ Based on a meta-analysis, most aversive effects were seen in repeated sessions lasting 60 min or more; therefore, we hypothesized that repetitive taVNS increased HRV but did not cause bradycardia and QT prolongation.^23^ To test this hypothesis, we compared changes in cardiovascular metrics at the phase of early brain injury (within 72 hours) and at the phase when delayed cerebral ischemia develops (after day 4) between patients receiving taVNS treatment and sham treatment. Root mean square of successive differences (RMSSD) and the standard deviation of normal RR intervals (SDNN) are two commonly used HRV metrics. RMSSD indicates parasympathetic activity, while lower SDNN is associated with increased cardiac risk.^27,28^ Providing effective taVNS treatment modulates the autonomic system, we propose that heart rate or HRV following acute taVNS could inform which SAH patients may experience the most clinical benefit from taVNS treatment. To explore this possibility, we correlated the changes in heart rate and HRV following acute taVNS treatment and changes in the modified Rankin Score (mRS), which measures the degree of disability or dependence in the daily activities of people suffering from neurological disability.

## Results

24 Participants were randomized to receive the taVNS (N = 11) or Sham (N = 13) treatment (**Table 1**, **Supplementary Figure 8)**. **Supplementary Table 3** shows the clinical characteristics of the two treatment groups. The participants, the medical team who dictated all management decisions for the patient’s subarachnoid hemorrhage, and the outcomes assessors who assigned modified Rankin Scores (mRS) at admission and discharge were blinded to the treatment. The structure of this study is shown in **Figure 1B**. Following randomization, enrolled patients underwent 20 minutes of either taVNS or sham stimulation twice daily during their stay in the ICU. This treatment schedule was informed by findings from Addorisio et al., where a 5-minute taVNS protocol was administered twice daily to patients with rheumatoid arthritis for two days.^29^ Their study found that circulating c-reactive protein (CRP) levels significantly reduced after 2 days of treatment but returned to baseline at the second clinical assessment by day 7. Given the high inflammatory state associated with SAH and our intention to maintain a steady reduction in inflammation, we decided to extend the treatment duration to 20 minutes per session. During treatment periods, a portable transcutaneous electrical nerve stimulation (TENS) device (TENS 7000 Digital TENS Unit, Compass Health Brands, OH, USA) was connected to the patient’s left ear using two ear clips (**Figure 1C and D**). For taVNS treatments, these ear clips were placed along the concha of the ear, while for sham treatments, the clips were placed along the earlobe to avoid stimulation of the auricular vagus nerve from tactile pressure (**Figure *1*C)**. For the taVNS group, stimulation parameters were selected based on values reported in prior studies that sought to maximize vagus somatosensory evoked potentials while avoiding the perception of pain: 20 Hz frequency, 250 µs pulse width, and 0.4 mA intensity.^30^ The stimulation parameters were designed to be imperceptible to the patients, and there were no reports of detection of taVNS, suggesting the success of the blinding. No electrical current was delivered during sham treatments. Please see ^32^ for a detailed protocol of this study.

**Table 1.** Patient demography. . HH: Hunt & Hess classification. mRS: modified Rankin Scale. Y: yes. N: no.

| Patient | Decade of life | Gender | Race | HH | mRS (admission) | mRS (discharge) | Vasospasm treated by blood pressure goal augmentation | Indwelling arterial lines | ICP monitoring |
| --- | --- | --- | --- | --- | --- | --- | --- | --- | --- |
| 1 | 70s | M | White | 3 | 5 | 5 | Y | Y | Y |
| 2 | 50s | M | Black/African American | 3 | 4 | 3 | N | Y | Y |
| 3 | 50s | F | White | 2 | 2 | 3 | N | N | N |
| 4 | 60s | M | White | 2 | 3 | 4 | Y | N | N |
| 5 | 60s | F | White | 2 | 2 | 3 | Y | N | N |
| 6 | 70s | F | White | 2 | 3 | 2 | N | Y | N |
| 7 | 60s | M | White | 4 | 5 | 4 | N | Y | Y |
| 8 | 60s | F | White | 4 | 5 | 2 | N | Y | Y |
| 9 | 70s | F | White | 2 | 3 | 0 | Y | N | N |
| 10 | 70s | F | White | 2 | 3 | 3 | Y | Y | N |
| 11 | 40s | M | Black/African American | 4 | 5 | 4 | Y | Y | Y |
| 12 | 40s | F | White | 2 | 4 | 3 | N | N | N |
| 13 | 50s | F | Black/African American | 2 | 2 | 1 | Y | N | N |
| 14 | 80s | F | White | 1 | 3 | 3 | Y | N | N |
| 15 | 70s | F | Black/African American | 4 | 5 | 3 | N | N | Y |
| 16 | 40s | F | Black/African American | 2 | 2 | 3 | N | N | N |
| 17 | 60s | F | White | 2 | 1 | 0 | N | N | N |
| 18 | 30s | F | White | 3 | 4 | 2 | N | N | Y |
| 19 | 60s | F | White | 3 | 4 | 2 | N | N | Y |
| 20 | 80s | F | White | 2 | 2 | 3 | N | Y | Y |
| 21 | 50s | F | White | 3 | 4 | 4 | N | Y | Y |
| 22 | 70s | F | White | 1 | 2 | 4 | N | N | N |
| 23 | 40s | F | Black/African American | 2 | 2 | 4 | N | Y | N |
| 24 | 60s | M | White | 4 | 5 | 5 | Y | Y | Y |

### Effects of repetitive taVNS on cardiac function

A study has shown that 15 minutes of taVNS reduced sympathetic activity in healthy individuals, with effects that persist during the recovery period.^33^ This finding suggests that taVNS may exert long-term effects on cardiovascular function. Therefore, we investigated whether repetitive taVNS treatment affects heart rate and QT interval, key indicators of bradycardia or QT prolongation, using 24-hour ECG recording. We found no significant differences in heart rate between groups (Mann–Whitney U test, N(taVNS) = 94, N(Sham)=95, p-value = 0.69, Cohen’s d =-0.01, W-statistics = 4317, power = 0.93). Changes in heart rate from Day 1 were equivalent between groups (Two-tailed equivalence tests, *p*[*lower threshold*] = 0.006, test statistics[*lower threshold*] = 2.53; *p*[*lower threshold*] = 0.004, test statistics[*lower threshold*] = -2.72, N(VNS)=94, N(VNS)=95). We further confirmed that changes in heart rate were similar between treatment groups following SAH (**Figure 3A** |Cohen’s d| < 0.2 for Day 2-4, Day 5-8, Day 8-10, and Day 11-13). Moreover, changes in corrected QT interval from Day 1 were significantly higher in the Sham group compared to the taVNS group (**Figure 3B**, Mann– Whitney U test, N(taVNS) = 94, N(Sham)=95, p-value < 0.001, Cohen’s d = -0.57). Similarly, uncorrected QT intervals from Day 1 were higher in the Sham group (**Supplementary Figure 10A**, Cohen’s d = -0.42). After the phase of early brain injury, the mean and median corrected QT interval were lower in the taVNS group with large effect sizes (**Figure 3B**, |Cohen’s d| > 0.5). To ensure that repetitive taVNS did not lead to QT prolongation outside the stimulation period, we calculated the percentage of prolonged QT intervals. Prolonged QT intervals were defined as corrected QT interval >= 500 ms. We found that changes in prolonged QT intervals percentage from Day 1 were higher in the Sham group (Figure 3F, Mann–Whitney U test, N(taVNS) = 94, N(Sham)=95, p-value < 0.001, Cohen’s d = -0.72).

Subsequently, we investigated the effect of taVNS treatment on RMSSD and SDNN. We found that changes in SDNN using Day 1 as baseline were not significantly different between the treatment groups (T-test, N(taVNS) = 94, N(Sham) = 95, p = 0.479, Cohen’s d =0.10, t statistics = 0.71, **Figure 2A**). Changes in RMSSD using Day 1 as baseline were significantly higher in the taVNS treatment group (T-test, N(taVNS) = 94, N(Sham) = 95, Bonferroni-corrected p = 0.025, Cohen’s d =0.42, t statistics = 2.91, **Figure 2B**). We further studied the effects of taVNS in different phases following SAH. **Figure 2A-B** show the changes in SDNN and RMSSD in bins of three days for the two treatment groups. The taVNS treatment increased RMSSD over the course of the treatment, with a smaller effect size (Cohen’s d = 0.29) observed between days 2-4, corresponding to the early brain injury phase, and large effect sizes at the later phases (Cohen’s d = 0.41 for Days 5–7, Cohen’s d = 0.54 for Days 8–10, Cohen’s d = 0.66 for Days 11–13). We further tested if the RMSSD reduction rate was greater in the Sham treatment group with a linear regression model: RMSSD change ∼ Day * Treatment. The results show that the RMSSD reduced slower in the taVNS treatment group when compared to the sham treatment, but this trend did not reach significance (coefficient of taVNS * Day interaction effect = 2.00, p = 0.21, **Supplementary Figure 1**).

**Figure 2.**
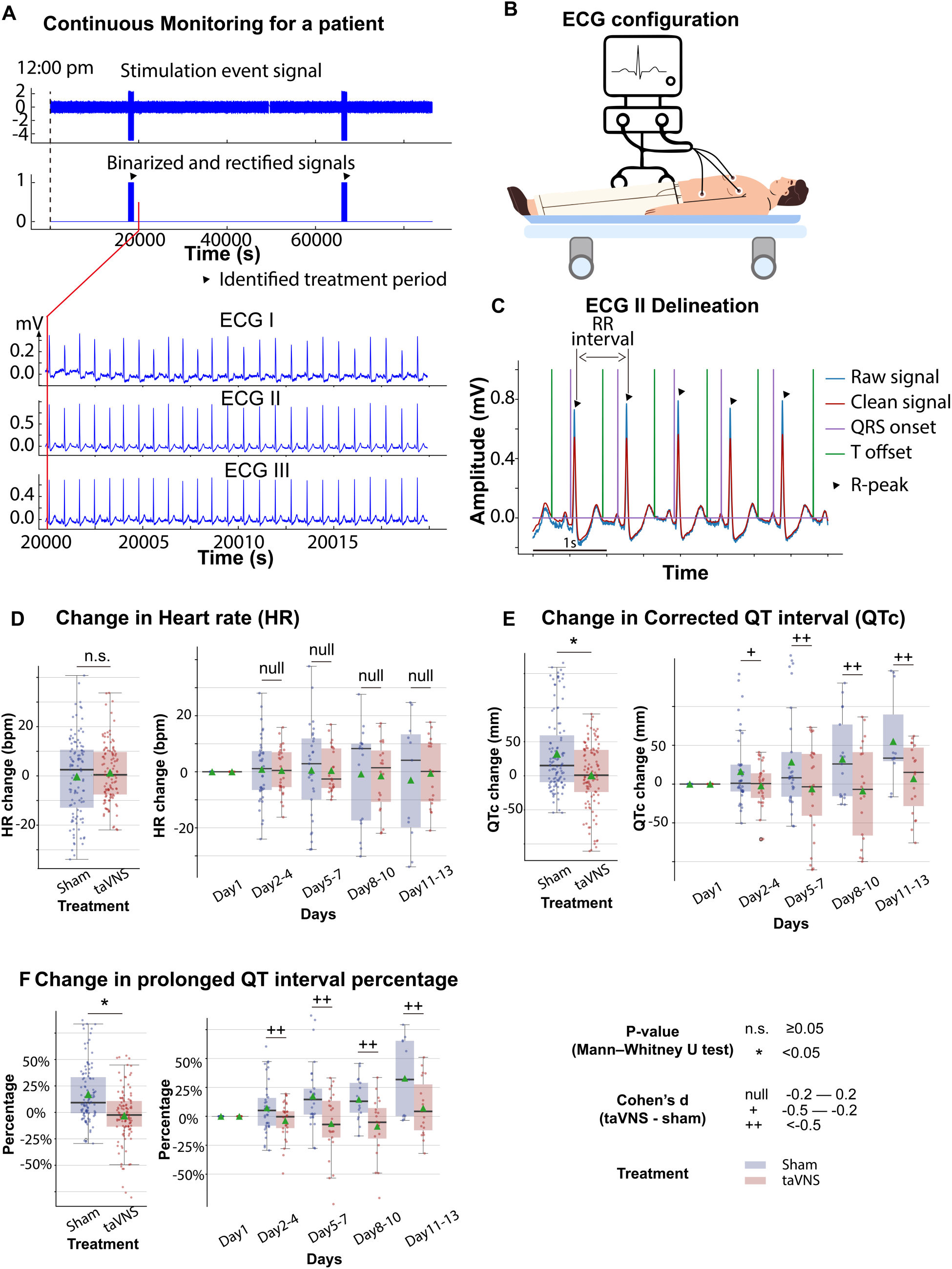
The effects of taVNS on cardiac function. **A**. Signals encoding treatment period and ECG signals in a representative patient. **B**. 3-lead ECG configuration in the intensive care unit. **C**. P wave, T wave, and QRS complex are delineated from clean ECG II signals. **D and E**. Heart rate and corrected QT interval changes from the first hospitalized day in the two treatment groups.

**Figure 3.**
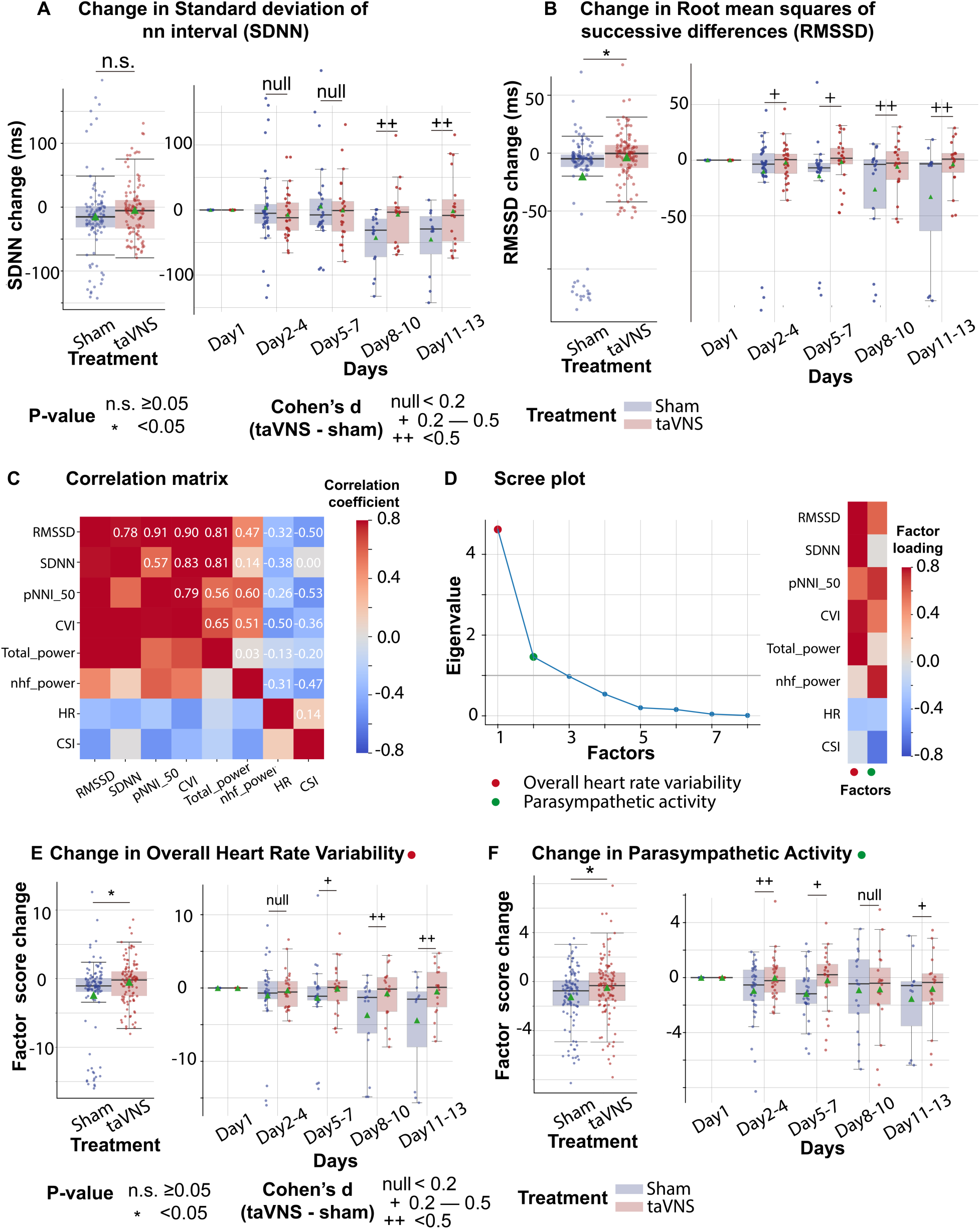
The effects of taVNS on overall heart rate variability and parasympathetic activity. **A-B**. Changes in standard deviation of NN interval (SDNN) changes and Root mean squares of successive differences over time for the two treatment groups. The color represents the treatment group. Green triangles represent the mean. **C**. Correlation between standard ECG features underlying autonomic activities. **D**. Factor analysis showed that there are two factors underlying the standard ECG features. The first factor is referred to as overall heart rate variability. The second factor is referred to as parasympathetic activity. **E-F**. The effect of taVNS on the two factors. pNNI_50: Percentage of Number of successive NN Intervals that differ by more than 50 ms. CVI: cardiac vagal index. Total power: total power below 0.4Hz of normal RR interval. nhf_power: relative power of the high-frequency band (0.15–0.4 Hz). CSI: cardiac sympathetic index.

RMSSD and SDNN are two of the most commonly used methods for quantifying heart rate variability. Bartlett’s test indicated that there are significant correlations among these measures (p < 0.01, **Figure 3C**). To analyze the effect of taVNS treatment on the autonomic system, we used factor analysis to identify the underlying factors. We focused on the two factors with the greatest eigenvalue. **Figure 3D** shows the factor loading, that is, the variance explained by heart rate variability metrics on the two factors. The first factor correlates positively with metrics representing variability, including RMSSD, SDNN, pNNI_50, and total power, and therefore has been termed Overall Heart Rate Variability. The second factor correlated positively with RMSSD and normalized high-frequency power, representing parasympathetic activity, and negatively correlated with the cardiac sympathetic index. Hence, it is termed Parasympathetic Activity. Overall Heart Rate Variability change from Day 1 was significantly higher in the VNS group (**Figure 3E**, Mann–Whitney U test, N(taVNS) = 94, N(Sham)=95, p-value = 0.04, Cohen’s d =0.37). The effect size was trivial between days 2-4 and increased over the course of treatment. The parasympathetic activity was also significantly higher in the VNS treatment group, and we observed the largest effect size between days 2-4 (Cohen’s d =0.50, **Figure 3F**).

We also investigated the potential association between clinical outcomes, as measured by changes in the mRS from admission to discharge, and heart rate variability metrics. We found that heart rate was lower in patients with improved mRS (i.e., <0) (Mann–Whitney U test, N(mRS < 0) = 122, N(mRS > 0)=98, p-value < 0.01, Cohen’s d =- 0.54). Parasympathetic Activity, Overall Heart Rate Variability, and corrected QT interval did not differ significantly between patients with improved mRS and patients with worsened mRS (**Supplementary Figure 4**).

### Effects of repetitive taVNS on vascular function

Elevated blood pressure is a common occurrence in SAH and is linked with a higher risk of re-rupture of cerebral aneurysms and vasospasm.^38,39^ In this study, patients in both treatment groups received medical treatment determined by the medical team, including vasopressors and medication for blood pressure management. We investigated whether taVNS induced any additional blood pressure changes beyond those managed by the medical team. We found that the median and mean blood pressure change from the first hospitalized day were greater than 0 for both treatment groups (**Figure *4*B**). No significant differences were detected in changes in blood pressure and intracranial pressure (ICP) between the treatment groups (**Figure *4*B and C)**. Equivalence testing confirmed that the ICP changes from the first hospitalization day were not significantly different between treatment groups, with a 2mmHg equivalence margin (two one-sided t-tests, *p*[*lower threshold*] = 3.66 x 10^-13^, t[*lower threshold*] = 8.07; *p*[*upper threshold*] = 3.33 x 10^-10^, t[*upper threshold*] = -6.73). Equivalence testing also indicated that there were no significant different changes in blood pressure between treatment groups (two one-sided t-tests, *p*[*lower threshold*] = 0.07, t[*lower threshold*] = 1.51; *p*[*upper threshold*] = 0.002, t[upper threshold] = -3.00). We further verified that there were no significant changes in arterial line blood pressure obtained via continuous invasive monitoring between treatment groups (**Supplementary Figure 5)**. We subsequently compared the Plethysmography Peripheral Perfusion Index (PPI) between the treatment groups as it is a proxy metric for cardiac stroke volume and vascular tone.^40,43^ We found that PPI change was significantly lower (Mann–Whitney U test, N(taVNS) = 83, N(Sham)=95, Bonferroni corrected p < 0.01, Cohen’s d =-0.49). In addition, respiration rate change was significantly higher (Mann–Whitney U test, N(taVNS) = 94, N(Sham)=95, Bonferroni corrected p = 0.02, Cohen’s d =0.37) in the taVNS group, as compared to the Sham group (**Figure *4*D and E**). We hypothesized that the increase in respiratory rate was a compensatory mechanism to ensure similar oxygen delivery. We found a significant negative correlation between changes in PPI and changes in respiration rate only for the taVNS treatment group (Pearson correlation coefficient = -0.37, p < 0.001, t-test, **Supplementary Figure 5D**). The Pearson correlation coefficient for the sham treatment group is -0.08 (p = 0.36).

**Figure 4.**
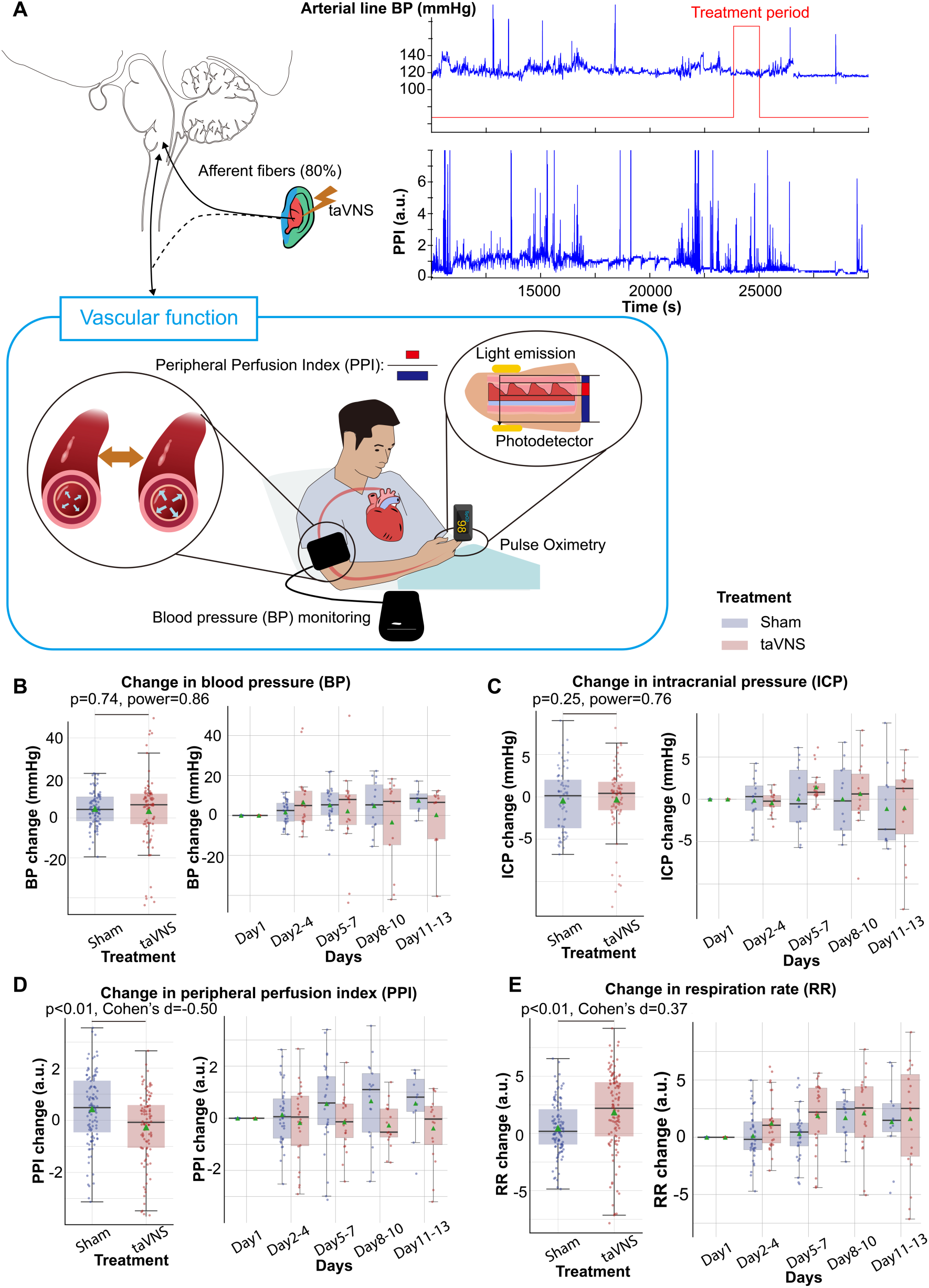
Effects of repetitive taVNS on vascular function. **A**. Representative vital signs and their physiology. Arterial line blood pressure (see supplementary Figure 5), intracranial pressure, and mean blood pressure measured regularly by nurses (BP) were recorded. Blood pressure is an index of vasodilation. PPI is the ratio between the pulsatile and the non-pulsatile blood flow, reflecting the cardiac output. **B-C.** Mean BP and ICP changes from the first hospitalization day did not differ significantly between the treatment groups. **D-E**. PPI change from the first hospitalized day was lower in the VNS treatment group, while RR change was higher.

### Acute effects of taVNS on cardiovascular function

Assessing the acute effect of taVNS on cardiovascular is crucial for its safe translation into clinical practice. We compared the acute change of heart rate, corrected QT interval, and heart rate variability between treatment groups, as abrupt changes in the pacing cycle may increase the risk of arrhythmias.^41^ The change in heart rate from treatment onset is shown in **Figure *5*B**. We subsequently tested whether taVNS affects changes in heart rate between post-treatment and pre-treatment. We found that the changes in heart rate were not significantly different between treatment groups although heart rate increased in the taVNS group (Wilcoxon rank-sum test, N = 188, Bonferroni corrected p = 0.03, Cohen’s d =0.11) but not in the Sham group (Wilcoxon signed ranked test, N = 199, Bonferroni corrected p = 0.72, Cohen’s d =0.00) (**Figure *5*C**). However, the increase in heart rate after taVNS was within 0.5 standard deviations of daily heart rate. There were no significant differences in changes in corrected QT interval or heart rate variability, as measured by RMSSD, SDNN, and relative power of high-frequency band between treatment groups (**Figure 5D and E** and **Supplementary Figure 6). Supplementary Figure 10B-C shows t**he acute changes in uncorrected QT interval. **Supplementary Table 3** summarizes the absolute changes in cardiovascular metrics for the treatment groups. We further asked whether heart rate can serve as a biomarker that indicates which SAH patients would receive the greatest benefit from continuing taVNS treatment. We investigated the relationship between changes in heart rate from pre- to post-taVNS treatment and changes in mRS between admission and discharge using a linear mixed-effects model. In this model, the treatment group, mRS change, and their interaction were included as fixed effects, while subject was included as a random effect. Our analysis revealed that the slope between changes in heart rate and changes in mRS is significantly more negative for the taVNS treatment group (**Table 2**). This finding suggests that an increase in heart rate following acute taVNS treatment is associated with improved clinical outcomes (**Figure *5*F**). Post-hoc analysis showed that patients in the taVNS treatment group who had an improvement in mRS of -2 or greater compared to other patients had significantly greater increases in heart rate (Mann-Whitney U test, p=0.02, N(mRS change <-1) = 53, N(mRS change >= -1) = 135, Cohen’s d=0.34, **Supplementary Figure 6C**). Conversely, HRV change, represented by RMSSD, was not significantly different based on mRS in SAH patients (**Supplementary Figure 6D**).

**Figure 5.**
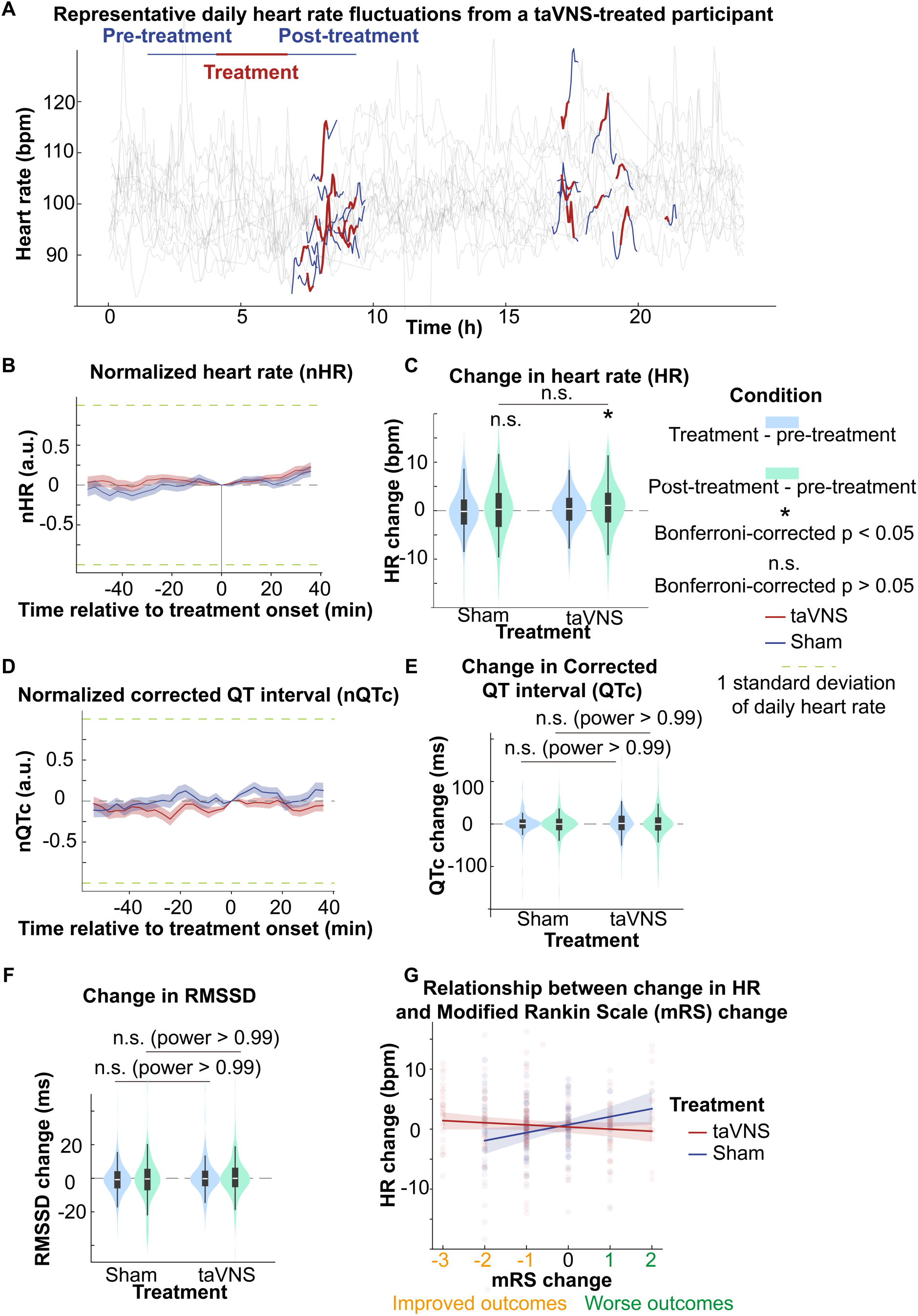
The acute effects of taVNS on cardiac function. **A**. Daily fluctuation of heart rate of a subject receiving VNS treatment. The treatment period, a 20-minute period before and after treatment, is highlighted. Note that a small proportion of ECG signals to derive heart rate was missing due to the expected cyclical restarting of the monitoring system. **B (D)**. Normalized heart rate (QTc) aligned at the treatment onset over time for the two treatment groups. The heart rate (QTc) is normalized based on the mean and standard error of heart rate for each day. **C**. The difference in HR between the treatment period, post-treatment period, and pre-treatment period for the two groups. Wilcoxon signed ranked test was used to test if the HR difference is statistically different from 0 in the VNS treatment group. Bonferroni-corrected p-value for HR difference between post-treatment and treatment period is 0.03 (N=188, Cohen’s d = 0.1). Mann–Whitney U tests were used to compare cardiac function metric differences between the two treatment groups. **E-F**. The difference in QTc and RMSSD between the treatment period, post-treatment period, and pre-treatment period for the two groups **G**. The relationship between heart rate changes following acute taVNS and functional outcome.

**Table 2.** Relationship between HR changes following acute taVNS and clinical outcomes.

| Model | $HR_{\text{subject,treatment}}(mRS) = \beta_0 + \beta_{\text{taVNS}} \times 1_{\text{treatment=taVNS}} + (\beta_{\text{mRS}} + \beta_{\text{taVNS,slope}} \times 1_{\text{treatment=taVNS}}) \times mRS + u_{\text{subject}}$ | | |
| --- | --- | --- | --- |
|  | Coefficient | P value | 95% Confidence interval |
| $\beta_0$ | 0.73 | 0.211 | -0.41 – 1.87 |
| $\beta_{\text{taVNS}}$ | -0.29 | 0.737 | -1.95 – 1.38 |
| $\beta_{\text{mRS}}$ | 1.47 | **0.006** | 0.43 – 2.51 |
| $\beta_{\text{taVNS,slope}}$ | -1.85 | **0.005** | -3.12 - 0.57 |

Subsequently, we compared changes in blood pressure, PPI, ICP, and respiration rate from pre- to post-treatment periods between treatment groups. We found that changes in PPI and blood pressure were significantly higher in the taVNS group, as compared to the Sham group (Mann-Whitney U test, blood pressure: *p* = 0.03, Cohen’s d = 0.22, N = 180 for Sham and 159 for taVNS; PPI: *p* < 0.01, Cohen’s d = 0.19, N = 227 for Sham and 186 for taVNS, **Supplementary Figure 7**). Only PPI remained significantly different between treatment groups after Bonferroni correction. The acute changes in PPI and blood pressure remained within the daily standard deviation. No significant differences in post-treatment changes in ICP or respiration rate were observed between treatment groups.

## Discussion

This study examined the effects of transcutaneous auricular vagus nerve stimulation (taVNS) on cardiovascular function in patients with subarachnoid hemorrhage (SAH). We investigated both the cumulative and acute impacts of taVNS. The findings in our study indicate that repetitive taVNS is not associated with previously suggested risks, such as bradycardia and QT prolongation. Furthermore, repetitive taVNS treatment increased overall heart rate variability and parasympathetic activity, which are indicators of a healthy cardiovascular system. When looking at the acute effects, taVNS only significantly increased the peripheral perfusion index but not heart rate, heart rate variability, corrected QT interval, blood pressure, or intracranial pressure. The findings are summarized in **Table 3**. Interestingly, we found that heart rate can serve as a biomarker for identifying SAH patients who are most likely to benefit from taVNS treatment. Collectively, this study substantiates the safety of treating SAH patients with taVNS and provides foundational data for future efforts to optimize and translate taVNS therapy toward clinical use.

**Table 3.** Summary of effects of acute and repetitive taVNS on cardiovascular function in SAH patients. . Metrics for cardiovascular function include heart rate variability, heart rate, QT interval, blood pressure, intracranial pressure, peripheral perfusion index, and respiration rate.

|  |  | Positive findings | Null findings | Implications |
| --- | --- | --- | --- | --- |
| Repetitive | Cardiac function | Increased HRV | QT interval<br>Heart rate | Increased parasympathetic activity |
|  | Vascular function | Reduced peripheral perfusion index<br>Increased respiration rate | Blood pressure<br>Intracranial pressure | Compensatory mechanisms to maintain autonomic balance |
| Acute | Cardiac function |  | Heart rate<br>Heart rate variability<br>QT interval | No to small acute effect |
|  | Vascular function | Increased peripheral perfusion index | Intracranial pressure<br>Respiration rate |  |

### taVNS and autonomic system

The autonomic nervous system (ANS), comprising the sympathetic and the parasympathetic nervous system, plays a critical role in maintaining physiological homeostasis. These two systems work synergistically to mediate interactions between the nervous and immune systems, which is thought to be the underlying mechanism for the immunomodulatory effect of taVNS. Our study is aligned with the finding that the autonomic balance is shifted toward sympathetic dominance following SAH (**Figure 3, Supplementary Figure 2**).^42,47^ In addition, we found that dysregulation of sympathovagal balance toward sympathetic dominance could be restored by taVNS treatment.

A key metric that reflects this restored sympathovagal balance is heart rate variability (**Figure 3F**). Specifically, factor analysis based on heart rate variability metrics showed that the parasympathetic activity was significantly higher in the taVNS treatment group. This difference was most pronounced during the early phase, between Days 2 and 4 following SAH. In addition to analyzing the correlation between the parasympathetic activity factor and established HRV measures that reflect parasympathetic activity, such as RMSSD and pNNI_50 (**Figure 3C**), we also examined changes in a frequency-domain HRV measure—the relative power of the high-frequency band (0.15–0.4 Hz)—to validate the accuracy of the factor analysis. The relative power of the high-frequency band is widely used to indicate respiratory sinus arrhythmia, a process primarily driven by the parasympathetic nervous system. We found that both the change in parasympathetic activity factor and relative high-frequency power were higher in the taVNS group at the early phase (Day 2 – 4, **Supplementary Figure 2**). Conversely, we observed higher high-frequency power in the Sham group during the later phase. If the factor analysis successfully isolates the parasympathetic activity, there should be other factors than the parasympathetic activity affecting the relative power of the high-frequency band. One such factor is the respiration rate. The high-frequency range is between 0.15 to 0.4 Hz, corresponding to respiration’s frequency range of approximately 9 to 24 breaths per minute. If the respiration rate increases and exceeds 24 breaths per minute, the respiratory-driven HRV might occur at a frequency higher than the typical high-frequency band. Given that the respiration rate was higher in the taVNS treatment group, a compensatory mechanism to ensure oxygen delivery (**Figure *4*E**), we hypothesized that the observed lower high-frequency power in the taVNS treatment group compared to sham at later phases was a result of increased respiration rate. Indeed, we found the normalized high-frequency power was higher when RR was less than 25 bpm compared to when RR > 25 bpm (Cohen’s d = 0.85, **Supplementary Figure 3A**). Moreover, an increase in RR in the taVNS treatment group was associated with a decrease in high-frequency power (**Supplementary Figure 3B**). These control analyses underscored the necessity of performing factor analysis to robustly measure parasympathetic activities and confirm that taVNS treatment mitigated the sympathetic overactivation during the early phase.

Additionally, taVNS led to a decreased QTc without a significant change in heart rate, mimicking the effects observed with propranolol administration, a beta-blocker that reduces sympathetic activity^53^. This finding suggests that repetitive taVNS reduces sympathetic overactivation and influences ventricular repolarization processes. Age affects sinus node function and is potentially associated with a higher risk of poor outcomes. To control for individual differences, including those due to age, our study compared the change in cardiovascular parameters from Day 1 within each subject across treatment groups. To further verify if age influences autonomic changes following SAH, we performed ANCOVA on autonomic function parameters with age included as a covariate. This analysis showed that age was negatively correlated with changes in heart rate, SDNN, and RMSSD from Day 1 but not with changes in QT intervals. After adjusting for age, we found that RMSSD and SDNN changes were significantly higher, while QTc changes were significantly lower in the taVNS treatment group (**Supplementary Table 4**). These results align with the conclusion that repetitive taVNS treatment increased HRV and was unlikely to cause bradycardia or QT prolongation. In addition, autonomic changes following SAH may be influenced by age. Specifically, lower RMSSD and SDNN in older individuals suggest a greater shift toward sympathetic predominance following SAH (**Supplementary Table 4**).

PPI is primarily influenced by cardiac output and vascular tone. Elevated PPI is associated with vasodilation and/or increased stroke volume. In the Sham group, increases in both PPI and blood pressure were observed when compared to Day 1 values (**Figure 3**). This effect may be due to higher stroke volume resulting from sympathetic activation following SAH. Alternatively, this could represent the heightened need for vasopressor interventions to improve cerebral perfusion due to more robust sympathetically driven cerebral vasospasm. The increase in PPI was less for the taVNS treatment group (**Figure *4***), suggesting a restored autonomic balance in the taVNS treatment group. However, the effects of taVNS on blood pressure require further investigation as more than half of the patients were on vasopressor and ionotropic drugs. Intuitively, sympathetic activation is associated with increases in both PPI and blood pressure. The blood pressure management might lead to similar blood pressure changes between the two treatment groups. Although repetitive taVNS increases heart rate variability days after initiation of the treatment, this effect is not seen acutely. Also, while repetitive taVNS was associated with a reduced PPI and no change in heart rate and blood pressure, there were small acute increases in PPI, heart rate, and blood pressure. All patients who were capable of verbal communication were asked if they felt any prickling or pain during all sessions. We confirmed that the current taVNS protocol is below the perception threshold for all trialed patients. Altogether, successful activation of the afferent vagal pathway by taVNS increased arousal, resulting in increased heart rate.^50,51^ These speculative mechanisms warrant further validation through animal or pharmacological studies directly investigating the effects of taVNS on autonomic function and vascular tone.

### Considerations for applying taVNS on SAH patients

Blood pressure management and cardiac function monitoring are crucial in patients following SAH.^38^ This study shows that blood pressure, QT interval, and heart rate over days were not significantly different between taVNS and sham treatment groups. This suggests that adding taVNS in treatments for SAH patients is unlikely to cause adverse blood pressure alterations or cardiac complications. Our findings suggest that repetitive taVNS could enhance parasympathetic tone following SAH. This effect could lead to favorable clinical outcomes as lower HRV was found to be associated with neurocardiogenic injury.^47,48^ Given the negative association between pro-inflammatory markers and HRV, our finding that HRV was higher in the taVNS treatment group aligns with the findings of primary outcomes of this clinical trial, which showed that taVNS treatment reduced pro-inflammatory cytokines, including tumor necrosis factor-alpha (TNF-α) and interleukin-6.^26,52^ The consistency between these findings strengthens the evidence supporting the anti-inflammatory effects of taVNS. In addition, the sympathetic predominance following SAH is implicated in an increased risk of delayed cerebral vasospasm, which is most commonly detected 5-7 days after SAH.^12^ Given that taVNS treatment mitigated the sympathetic overactivation before the typical onset of cerebral vasospasm, it could potentially reduce the severity of this complication. Additionally, reduced PPI was associated with increased respiration rate only for the taVNS treatment group, suggesting that the autonomic system self-regulates to maintain cardiovascular homeostasis. Thus, it is important to consider autonomic system self-regulation when studying the therapeutic effects of taVNS.^44^ Also, while acute cardiovascular changes were noted after taVNS, these changes were within normal daily variations in this study, making them unlikely to pose a risk to the patient. That said, the observed acute increases in PPI following taVNS necessitate caution when considering taVNS treatment for patients to whom peripheral vasodilatation is not desired.

As we pioneer the application of taVNS as an immunomodulation technique in SAH patients, we adopt parameters (20 Hz, 0.4 mA) reported in similar studies.^55^ The current study provides a basis for future preclinical and clinical studies of taVNS in this patient population. To build on our findings, a systematic evaluation of the relationship between parameters such as frequency, intensity, and duration and taVNS’s effects on the immune system and cardiovascular function is necessary to establish taVNS as an effective therapeutic option for SAH patients.^56^

## Limitations and outlook

While this study supports the safety of taVNS treatment in SAH patients, we should be cautious when generalizing these findings to broader clinical populations. The current study did not explore the effects of taVNS on less commonly used cardiovascular metrics, such as QTc dispersion. Our study considers each day as an independent sample for the following considerations: 1. heart rate and HRV metrics exhibited great daily variations. Their value on one day was not predictive of the metrics on another day, which could be due to medications, interventions, or individualized SAH recovery process during the patient’s stay in the ICU. 2. SAH patients in the ICU often experience daily changes in clinical status, including fluctuations in intracranial pressure, blood pressure, neurological status, and other vital signs. 3. Day-to-day cardiovascular function changes varied as the patient recovered or encountered setbacks. To conclusively establish that there is no significant cardiovascular effect of repetitive taVNS on any given day following SAH, we would need to perform statistical tests between treatment groups for each day. In this context, 64 subjects per treatment group are required to achieve 80% power, assuming a medium effect size (Cohen ‘s d = 0.5) and 0.05 type I error probability (two-sample t-test).

Mild cardiac abnormalities are common in SAH patients^5^, complicating the precise calculation of cardiovascular metrics from ECG signals and the interpretation of the results. Systematic verification of methods for calculating cardiovascular metrics to ensure their applicability in SAH patients is crucial. We noticed a high variance of change in heart rate for days 5 – 7, 8 – 10, and 11 – 13 for both treatment groups (**Figure 2D**). This may be due to the small sample size in the later days, given that the mean duration of hospitalization for the 24 subjects included in this study was 11.3 days with a standard deviation of 6.4. Differences in medical history and clinical outcomes during hospitalization may also explain the variance of change in heart rate for the later days. For example. heart rate was lower in patients with improved mRS scores (Supplementary Figure 4B). Understanding the association between cardiovascular metrics and clinical assessments, such as vasospasm and inflammation, could help decide whether future taVNS trials should control for these factors when evaluating the effects of taVNS on cardiovascular function. Additional care should be paid when interpreting the results of blood pressure, as hypertension was intentionally induced for some patients being treated for vasospasm. Patient medical histories are summarized in **Table 1**.

Kulkarni et al. showed that the response to low-level tragus stimulation (LLTS) varied among patients with atrial fibrillation.^54^ Similarly, in our study, not all patients in the taVNS treatment group showed a reduction in mRS scores (improved degree of disability or dependence). This differential response may be inherent to taVNS and potentially influenced by factors such as anatomical variations in the distribution of the vagus nerve at the outer ear. These findings underscore the importance of using acute biomarkers to guide patient selection and optimize stimulation parameters. Furthermore, we found that increased heart rate was a potential acute biomarker for identifying SAH patients who are most likely to respond favorably to taVNS treatment. Translating this finding into clinical practice will require further research to elucidate the mechanisms by which an acute increase in heart rate may predict the outcomes of patients receiving taVNS, including its relationship with neurological evaluations, vasospasm, echocardiography, and inflammatory markers.

## Conclusions

Utilizing taVNS as a neuromodulation technique in SAH patients is safe without inducing bradycardia or QT prolongation. Repetitive taVNS treatment increased parasympathetic activity. Acute taVNS elevated heart rate, which might be an acute biomarker to identify SAH patients who are likely to respond favorably to taVNS treatment.

## Methods details

### Study Participants

Participants in this study were recruited from adult patients who were admitted to the ICU at Barnes Jewish Hospital, St. Louis, MO, following an acute, spontaneous, aneurysmal SAH. Inclusion criteria were: (**1**) Patients with SAH confirmed by CT scan; (**2**) Age > 18; (**3**) Patients or their legally authorized representative are able to give consent. Exclusion criteria were: (**1**) Age < 18; (**2**) Use of immunosuppressive medications; (**3**) Receiving ongoing cancer therapy; (**4**) Implanted electrical device; (**5**) Sustained bradycardia on admission with a heart rate < 50 beats per minute for > 5 minutes; (**6**) Considered moribund/at risk of imminent death. Participants were randomized to receive either the taVNS (N = 11) or Sham (N = 13) treatment. Patients were enrolled prior to randomization by a member of the research team who went through the informed consent process with the patient or their legally authorized representative. Treatment group assignment was via a computer-generated randomization sequence, with the next number obscured until patient enrollment. Research team members who applied the ear clips and set stimulation parameters were not blinded to the treatment. The participants, the medical team who dictated all management decisions for the patient’s subarachnoid hemorrhage, and the outcomes assessors who assigned modified Rankin Scores (mRS) at admission and discharge were blinded to the treatment. The structure of this study is shown in **Figure 1B**. This study was approved by the Washington University School of Medicine Review Board and was conducted in accordance with institutional and national ethics guidelines and the Declaration of Helsinki (Clinical trial number: NCT04557618).

### taVNS protocol

Following randomization, enrolled patients underwent 20 minutes of either taVNS or sham stimulation twice daily during their stay in the ICU. During treatment periods, a portable transcutaneous electrical nerve stimulation (TENS) device (TENS 7000 Digital TENS Unit, Compass Health Brands, OH, USA) was connected to the patient’s left ear using two ear clips (**Figure 1C and D**). For taVNS treatments, these ear clips were placed along the concha of the ear, while for sham treatments, the clips were placed along the earlobe to avoid stimulation of the auricular vagus nerve from tactile pressure (**Figure 1 Figure 1C)**. For the taVNS group, stimulation parameters were selected based on values reported in prior studies that sought to maximize vagus somatosensory evoked potentials while avoiding the perception of pain: 20 Hz frequency, 250 µs pulse width, and 0.4 mA intensity^30^. The stimulation was not perceptible for the patients. No electrical current was delivered during sham treatments. For both groups, the TENS device was connected to the patient and a bedside recording computer. The computer recorded continuous ECG and vital signs, including blood pressure, temperature, respiration rate, peripheral perfusion index, intracranial pressure, and arterial blood pressure. The collection of intracranial pressure and arterial blood pressure data varied, being dependent on the treatment protocol assigned by the clinical team, and thus was not uniformly available for all patients throughout the study. Please see ^32^ for a detailed protocol of this study.

### Data processing

A 3-lead system was used for electrocardiograms (ECG). ECG signals, sampled at 500 Hz, and other vital signs, such as blood pressure, sampled at 1 Hz, were recorded from the Intellivue patient monitor (Philips®, Netherlands) using vitalDB software.^34^

To calculate cardiac metrics, we first applied a 0.5 Hz fifth-order high-pass Butterworth filter and a 60 Hz powerline filter on ECG data to reduce artifacts. ^35^ We detected QRS complexes based on the steepness of the absolute gradient of the ECG signal using the Neurokit2 software package.^35^ R-peaks were detected as local maxima in the QRS complexes. P waves, T waves, and QRS complexes were delineated based on the wavelet transform of the ECG signals proposed by Martinez J. P. et al. (**Figure 2A-C**).^36^ This algorithm identifies the QRS complex by searching for modulus maxima, which are peaks in the wavelet transform coefficients that exceed specific thresholds. The onset of the QRS complex is determined as the beginning of the first modulus maximum before the modulus maximum pair created by the R wave. To identify the T wave, the algorithm searches for local maxima in the absolute wavelet transform in a search window defined relative to the QRS complex. Thresholding is used to identify the offset of the T wave. RR intervals were preprocessed to exclude outliers, defined as RR intervals greater than 2 s or less than 300 ms. RR intervals with > 20% relative difference to the previous interval were considered ectopic beats and excluded from analyses. After preprocessing, RR intervals were used to calculate heart rate, heart rate variability, and corrected QT (QTc) based on Bazett’s formula: 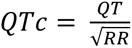.^46^ The corrected QT interval (QTc) estimates the QT interval at a standard heart rate of 60 bpm. Heart rate variability measures included the root mean square of successive difference of normal RR intervals (RMSSD), indicating parasympathetic activity, and the standard deviation of normal RR intervals (SDNN), a clinical measure of cardiac risk.^27,28^ Heart rate variability calculations are detailed in **Supplementary Materials**.

To investigate the effect of repetitive taVNS on cardiovascular function, we compared heart rate variability, heart rate, corrected QT intervals, blood pressure, and intracranial pressure calculated over 24 hours between patients receiving taVNS and sham treatment. In addition, we compared the mean peripheral perfusion index and respiration rate over 24 hours between treatment groups to determine the effects of repetitive taVNS on the autonomic system. Data collection commenced on the first day of each patient’s ICU admission. The average duration of continuous data recording was 11.1 days, with a standard deviation of 6.8 days. To analyze the effects of taVNS treatment more granularly, we segmented the changes in these metrics from the initial day at three-day intervals, facilitating comparison between the taVNS and sham treatment groups over the course of their ICU stay.

To study the effects of acute treatment over time, we focused on blood pressure, heart rate variability, heart rate, and corrected QT intervals 20 minutes before treatment (pre-treatment), during the 20-minute treatment (during-treatment), and 20 minutes after treatment (post-treatment). The treatment event signals were rectified and binarized based on their half-maximum value to identify the treatment onset and offset (**Figure 2A**). We calculated metrics using 6-minute sliding windows over ECG data starting from treatment onset/offset and moving bi-directionally with a 3-minute step. To correct daily and between-subject variation, we applied the same sliding window strategy to calculate the mean and standard deviation of these cardiac metrics for each patient each day as a reference. Subsequently, heart rate variability, heart rate, and corrected QT interval around treatment onset/offset were normalized based on the reference. In addition, we calculated the difference in blood pressure, heart rate variability, and heart rate, and corrected QT intervals between during-treatment and pre-treatment, as well as the difference between post-treatment and pre-treatment for each patient and for each treatment. To study the effects of acute taVNS, we compared the two differences between the treatment groups.

### Factor Analysis

We performed an exploratory factor analysis to identify the factors underlying autonomic system activity. Besides RMSSD and SDNN, variables derived from preprocessed RR intervals and used to perform factor analysis included the percentage of successive normal-to-normal (NN) Intervals that differ by more than 50 ms (pNNI_50), total power (below 0.4 Hz), normalized high-frequency power (0.15-0.4Hz), cardiac vagal index, and cardiac sympathetic index. The total power is thought to represent the overall heart rate variability, while normalized high-frequency power primarily reflects parasympathetic activity.^27^ These variables were normalized using a z-score method based on individual daily means and standard deviations before factor analysis. Factor analysis was performed using the factor_analyzer Python package. The number of factors was set to 2 based on the Scree plot. The factor loadings were calculated using the Minimum Residual Method. After factor extraction, a Varimax rotation was applied for better interpretability so that each factor had high loadings for a smaller number of variables and low loadings for the remaining variables.

### Statistical Analyses

To investigate the effect of taVNS at the phase of early brain injury and later phases, we grouped the change of heart rate variability, heart rate, and corrected QT interval from the first hospitalized day in bins of three days. The change in blood pressure, intracranial pressure, respiration rate, and peripheral perfusion index from the first hospitalization day were also compared between treatment groups. We used t-tests for comparisons between treatment groups when the data were normally distributed, as determined by the Shapiro-Wilk test. We employed Mann–Whitney U tests for non-normally distributed data. We used Wilcoxon signed-rank tests to compare the difference in heart rate between post-treatment and during-treatment against 0. To control the familywise error rate, we applied Bonferroni correction. Specifically, when investigating the cardiac effects of taVNS, we compared six metrics between treatment groups, including heart rate, corrected QT interval, RMSSD, SDNN, and two factors representing heart rate variability. Consequently, the p-values were corrected by a factor of six. In this study, we reported the statistical power achieved for tests that yielded non-significant results. The achieved power is calculated based on a two-sample t-test assuming a medium effect size (Cohen’s d of 0.5) and a Type I error probability (a) of 0.05. We used two one-sided tests to confirm that taVNS did not induce long-term changes in heart rate, corrected QT interval, or blood pressure, with equivalency test margins set to 5 bpm for heart rate, 50 ms for QT interval, and 2 mmHg for blood pressure. A summary of statistical tests is provided in **Supplementary Table 1**.

## Data Availability

All data produced in the present study are available upon reasonable request to the authors

## Availability of data and materials

https://github.com/GanshengT/taVNS_SAH

## Authors’ contributions

**Gansheng Tan**: Conceptualization, Methodology, Investigation, Formal analysis, Software, Writing – original draft, Writing – review & editing. **Anna L. Huguenard**: Conceptualization, Funding acquisition, Methodology, Data curation, Writing - original draft, Writing – review & editing. **Kara M. Donovan**: Writing – review & editing. **Philip Demarest**: Writing – review & editing. **Xiaoxuan Liu**: Methodology, Writing – review & editing. **Ziwei Li**: Methodology, Writing – review & editing. **Markus Adamek**: Methodology, Software. **Kory Lavine**: Writing – review & editing. **Ananth K. Vellimana**: Data curation, Writing – review & editing. **Terrance T. Kummer:** Data curation, Writing – review & editing. **Joshua W. Osbun**: Data curation, Writing – review & editing. **Gregory J. Zipfel**: Funding acquisition, Resources, Writing - review & editing. **Peter Brunner**: Funding acquisition, Resources, Supervision, Writing - review & editing. **Eric C. Leuthardt**: Conceptualization, Supervision, Funding acquisition, Writing – review & editing, Writing – original draft.

## Acknowledgments

The authors acknowledge physicians/nurses for helping administer treatment. The authors thank Dr. Paul Cassidy for his contributions to the scientific editing of this manuscript, supported by the Institute of Clinical and Translational Sciences grant UL1TR002345 from the National Center for Advancing Translational Sciences (NCATS).

## References

1. D’Souza, S. (2015). Aneurysmal Subarachnoid Hemorrhage. In Journal of Neurosurgical Anesthesiology (Vol. 27, Issue 3, pp. 222–240). Ovid Technologies (Wolters Kluwer Health).

2. Provencio, J. J. Inflammation in Subarachnoid Hemorrhage and Delayed Deterioration Associated with Vasospasm: A Review. Acta Neurochirurgica Supplement 233–238 (2012)

3. Wu, Z. et al. Transcutaneous auricular vagus nerve stimulation reduces cytokine production in sepsis: An open double-blind, sham-controlled, pilot study. Brain Stimulation vol. 16 507–514 (2023).

4. Schneider, U. C., Xu, R. & Vajkoczy, P. Inflammatory Events Following Subarachnoid Hemorrhage (SAH). Current Neuropharmacology vol. 16 1385–1395 (2018).

5. Norberg, E., Odenstedt-Herges, H., Rydenhag, B. & Oras, J. Impact of Acute Cardiac Complications After Subarachnoid Hemorrhage on Long-Term Mortality and Cardiovascular Events. Neurocritical Care vol. 29 404–412 (2018).

6. Weir, B. Unruptured intracranial aneurysms: a review. Journal of Neurosurgery vol. 96 3–42 (2002).

7. van Gijn, J., Kerr, R. S. & Rinkel, G. J. Subarachnoid hemorrhage. The Lancet vol. 369 306–318 (2007).

8. Tracey, K. J. The inflammatory reflex. Nature vol. 420 853–859 (2002).

9. Lv, S. et al. Levels of Interleukin-1β, Interleukin-18, and Tumor Necrosis Factor-α in Cerebrospinal Fluid of Aneurysmal Subarachnoid Hemorrhage Patients May Be Predictors of Early Brain Injury and Clinical Prognosis. World Neurosurgery vol. 111 e362–e373 (2018).

10. Bonaz, B. et al. Chronic vagus nerve stimulation in Crohn’s disease: a 6-month follow-up pilot study. Neurogastroenterology & Motility vol. 28 948–953 (2016).

11. Macdonald, R. L. et al. Randomized Trial of Clazosentan in Patients With Aneurysmal Subarachnoid Hemorrhage Undergoing Endovascular Coiling. Stroke vol. 43 1463– 1469 (2012).

12. Budohoski, K., Czosnyka, M., Kirkpatrick, P. et al. Clinical relevance of cerebral autoregulation following subarachnoid haemorrhage. Nat Rev Neurol 9, 152–163 (2013).

13. Meyers, E. C. et al. Vagus Nerve Stimulation Enhances Stable Plasticity and Generalization of Stroke Recovery. Stroke vol. 49 710–717 (2018).

14. Provencio, J. J. & Vora, N. Subarachnoid Hemorrhage and Inflammation: Bench to Bedside and Back. Seminars in Neurology vol. 25 435–444 (2005).

15. Huguenard A, Tan G, Johnson G, et al O-055 Non-invasive auricular vagus nerve stimulation following spontaneous subarachnoid hemorrhage reduces rates of radiographic vasospasm and hospital-acquired infections Journal of NeuroInterventional Surgery 2023;15:A43–A44.

16. Frangos, E., Ellrich, J. & Komisaruk, B. R. Non-invasive Access to the Vagus Nerve Central Projections via Electrical Stimulation of the External Ear: fMRI Evidence in Humans. Brain Stimulation vol. 8 624–636 (2015).

17. Sahn, B., Pascuma, K., Kohn, N., Tracey, K. J. & Markowitz, J. F. Transcutaneous auricular vagus nerve stimulation attenuates inflammatory bowel disease in children: a proof-of-concept clinical trial. Bioelectronic Medicine vol. 9 (2023).

18. Pavlov, V. A. & Tracey, K. J. The vagus nerve and the inflammatory reflex—linking immunity and metabolism. Nature Reviews Endocrinology vol. 8 743–754 (2012).

19. Tynan, A., Brines, M. & Chavan, S. S. Control of inflammation using non-invasive neuromodulation: past, present and promise. International Immunology vol. 34 119– 128 (2021).

20. Keute, M., Machetanz, K., Berelidze, L., Guggenberger, R. & Gharabaghi, A. Neuro-cardiac coupling predicts transcutaneous auricular vagus nerve stimulation effects. Brain Stimulation vol. 14 209–216 (2021).

21. Naggar, I. et al. Vagal control of cardiac electrical activity and wall motion during ventricular fibrillation in large animals. Autonomic Neuroscience vol. 183 12–22 (2014).

22. Hua, K. et al. Cardiovascular effects of auricular stimulation -a systematic review and meta-analysis of randomized controlled clinical trials. Frontiers in Neuroscience vol. 17 (2023).

23. Kim, A. Y. et al. Safety of transcutaneous auricular vagus nerve stimulation (taVNS): a systematic review and meta-analysis. Scientific Reports vol. 12 (2022).

24. Schmidt, J. M. et al. Prolonged Elevated Heart Rate is a Risk Factor for Adverse Cardiac Events and Poor Outcome after Subarachnoid Hemorrhage. Neurocritical Care vol. 20 390–398 (2013).

25. Zhang, L. & Qi, S. Electrocardiographic Abnormalities Predict Adverse Clinical Outcomes in Patients with Subarachnoid Hemorrhage. Journal of Stroke and Cerebrovascular Diseases vol. 25 2653–2659 (2016).

26. Huguenard, A. L. et al. Auricular Vagus Nerve Stimulation Mitigates Inflammation and Vasospasm in Subarachnoid Hemorrhage: A Randomized Trial. (2024) doi:10.1101/2024.04.29.24306598.

27. Kleiger, R. E., Stein, P. K. & Bigger, J. T., Jr. Heart Rate Variability: Measurement and Clinical Utility. Annals of Noninvasive Electrocardiology vol. 10 88–101 (2005).

28. Shaffer, F. & Ginsberg, J. P. An Overview of Heart Rate Variability Metrics and Norms. Frontiers in Public Health vol. 5 (2017).

29. Addorisio, M. E. et al. Investigational treatment of rheumatoid arthritis with a vibrotactile device applied to the external ear. Bioelectronic Medicine vol. 5 (2019).

30. de Gurtubay, I. G., Bermejo, P., Lopez, M., Larraya, I. & Librero, J. Evaluation of different vagus nerve stimulation anatomical targets in the ear by vagus evoked potential responses. Brain and Behavior vol. 11 (2021).

31. van der Bilt, I. A. C. et al. Impact of cardiac complications on outcome after aneurysmal subarachnoid hemorrhage. Neurology vol. 72 635–642 (2009).

32. Huguenard, A. et al. Non-invasive Auricular Vagus nerve stimulation for Subarachnoid Hemorrhage (NAVSaH): Protocol for a prospective, triple-blinded, randomized controlled trial. PLOS ONE vol. 19 e0301154 (2024).

33. Clancy, J. A. et al. Non-invasive Vagus Nerve Stimulation in Healthy Humans Reduces Sympathetic Nerve Activity. Brain Stimulation vol. 7 871–877 (2014).

34. Lee, H.-C. & Jung, C.-W. Vital Recorder—a free research tool for automatic recording of high-resolution time-synchronised physiological data from multiple anaesthesia devices. Scientific Reports vol. 8 (2018).

35. Schölzel, C., & Chen, S. A. (2021). NeuroKit2: A Python toolbox for neurophysiological signal processing. Behavior Research Methods, 53(4), 1689–1696. 10.3758/s13428-020-01516-y

36. Martinez, J. P., Almeida, R., Olmos, S., Rocha, A. P. & Laguna, P. A Wavelet-Based ECG Delineator: Evaluation on Standard Databases. IEEE Transactions on Biomedical Engineering vol. 51 570–581 (2004).

37. Naredi, S. et al. Increased Sympathetic Nervous Activity in Patients With Nontraumatic Subarachnoid Hemorrhage. Stroke vol. 31 901–906 (2000).

38. Minhas, J. S., Moullaali, T. J., Rinkel, G. J. E. & Anderson, C. S. Blood Pressure Management After Intracerebral and Subarachnoid Hemorrhage: The Knowns and Known Unknowns. Stroke vol. 53 1065–1073 (2022).

39. Hosmann, A. et al. Endogenous arterial blood pressure increase after aneurysmal subarachnoid hemorrhage. Clinical Neurology and Neurosurgery vol. 190 105639 (2020).

40. Elshal, M. M., Hasanin, A. M., Mostafa, M. & Gamal, R. M. Plethysmographic Peripheral Perfusion Index: Could It Be a New Vital Sign? Frontiers in Medicine vol. 8 (2021).

41. Zaniboni, M. The electrical restitution of the non-propagated cardiac ventricular action potential. Pflügers Archiv - European Journal of Physiology vol. 476 9–37 (2023).

42. Chiu, T.-F., Huang, C.-C., Chen, J.-H. & Chen, W.-L. Depressed sympathovagal balance predicts mortality in patients with subarachnoid hemorrhage. The American Journal of Emergency Medicine vol. 30 651–656 (2012).

43. Coutrot, M. et al. Perfusion index: Physical principles, physiological meanings and clinical implications in anaesthesia and critical care. Anaesthesia Critical Care & Pain Medicine vol. 40 100964 (2021).

44. Bjerkne Wenneberg, S., et al. Heart rate variability monitoring for the detection of delayed cerebral ischemia after aneurysmal subarachnoid hemorrhage. Acta Anaesthesiologica Scandinavica vol. 64 945–952 (2020).

45. Zhang, A. et al. Clinical Potential of Immunotherapies in Subarachnoid Hemorrhage Treatment: Mechanistic Dissection of Innate and Adaptive Immune Responses. Aging and disease vol. 14 1533 (2023).

46. Bazett, H.C. (1920) An Analysis of the Time-Relations of Electrocardiograms. Heart, 7, 353–370.

47. Bai, X. et al. The Clinical Characteristics of Heart Rate Variability After Stroke. The Neurologist vol. 29 133–141 (2023).

48. Megjhani, M. et al. Heart Rate Variability as a Biomarker of Neurocardiogenic Injury After Subarachnoid Hemorrhage. Neurocritical Care vol. 32 162–171 (2019).

49. Sharon, O., Fahoum, F. & Nir, Y. Transcutaneous Vagus Nerve Stimulation in Humans Induces Pupil Dilation and Attenuates Alpha Oscillations. The Journal of Neuroscience vol. 41 320–330 (2020).

50. Skora, L., Marzecová, A. & Jocham, G. Tonic and phasic transcutaneous auricular vagus nerve stimulation (taVNS) both evoke rapid and transient pupil dilation. Brain Stimulation vol. 17 233–244 (2024).

51. Tan, G. et al. Does vibrotactile stimulation of the auricular vagus nerve enhance working memory? A behavioral and physiological investigation. Brain Stimulation vol. 17 460–468 (2024).

52. Williams, D. P. et al. Heart rate variability and inflammation: A meta-analysis of human studies. Brain, Behavior, and Immunity vol. 80 219–226 (2019).

53. Solti, F., Szatmáry, L., Vecsey, T. & Szabolcs, Z. The effect of sympathetic and parasympathetic activity on QT duration. Clinical study in patients with normal and prolonged QT time. Cor Vasa 31, 9–15 (1989).

54. Kulkarni, K. et al. Low-Level Tragus Stimulation Modulates Atrial Alternans and Fibrillation Burden in Patients With Paroxysmal Atrial Fibrillation. Journal of the American Heart Association vol. 10 (2021).

55. Jelinek, M., Lipkova, J. & Duris, K. Vagus nerve stimulation as immunomodulatory therapy for stroke: A comprehensive review. Experimental Neurology vol. 372 114628 (2024).

56. Dusi, V., Angelini, F., Zile, M. R. & De Ferrari, G. M. Neuromodulation devices for heart failure. European Heart Journal Supplements vol. 24 E12–E27 (2022)

