## Supplementary Figure 1 for "The effect of transcutaneous auricular vagus nerve stimulation on cardiovascular function in subarachnoid hemorrhage patients: a safety study"

**Calculation of cardiovascular metrics**

1. Heart rate (HR) over a period is the mean of $\frac{60}{NN interval in seconds}$ over this period.
2. The percentage of prolonged QT intervals is defined as the number of intervals with a corrected QT interval (QTc) ≥ 500 ms, divided by the total number of QT intervals.
3. Root mean square of successive difference of normal RR intervals (RMSSD) for a given period:

$RMSSD =\sqrt{\frac{1}{N-1}\sum_{i}^{N-1} {({NN}_{i+1}-{NN}_{i})}^{2}}$, where N is the number of NN intervals within this period.

4. Standard deviation of normal RR intervals (SDNN) for a given period:

$SDNN =\sqrt{\frac{1}{N}\sum_{i}^{N} {({NN}_{i}-\bar{NN})}^{2}}$, where $\bar{NN}$ represents the mean of NN intervals within this period.

5. To calculate total power and normalized high-frequency power, power spectral density is calculated from NN intervals with the help of Python package *hrv-analysis*. Linear interpolation is used to make NN intervals evenly spaced, assuming a sampling frequency of 4Hz. Welch’s method is used to calculate power spectral density $P(f)$.

$$Total power = \int_{0}^{0.4} P(f) df$$

$$High-frequency power = \int_{0.15}^{0.4} P(f) df$$

$$Normalized high-frequency power =\frac{High-frequency power}{Total power}$$

6. Cardiac vagal index (CVI) and cardiac sympathetic index (CSI):

The width (SD1) and length (SD2) of the Poincaré plot, a graphical representation of the correlation between consecutive NN intervals, are used to calculate CVI and CSI.

$$CSI = \frac{SD1}{SD2}$$

$$CVI = {log}_{10}(SD1 * SD2)$$

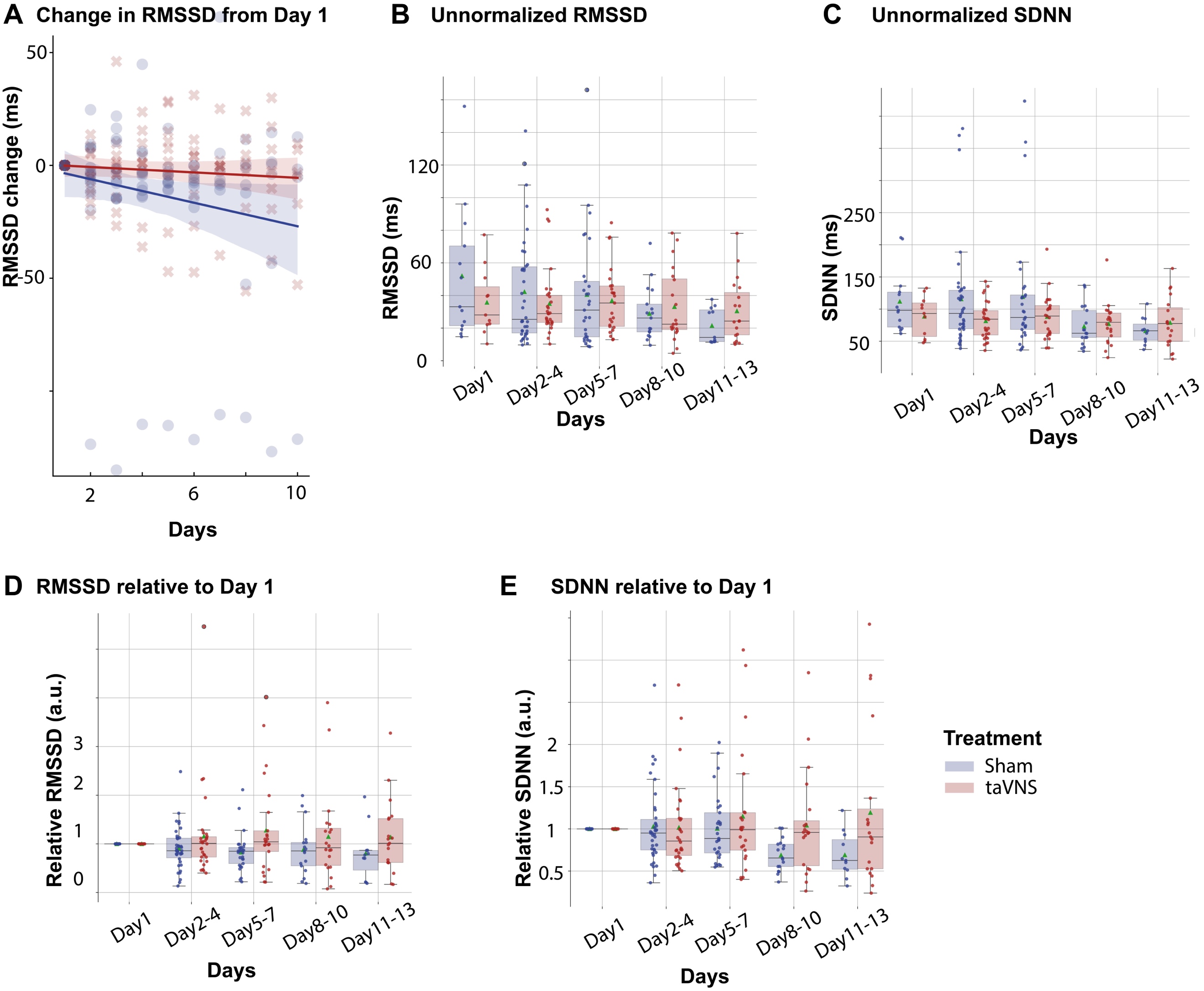

**Supplementary Figure 1** (related to Figure 2) **Heart rate variability in taVNS and Sham treatment groups. A.** In the Linear regression model (RMSSD change ~ Day * Treatment), the coefficient for interaction effect is 2.01 (p = 0.21), the coefficient for Day is -2.61 (p=0.02), and the coefficient for Treatment is 1.38 (p = 0.88). RMSSD: Root mean square of successive differences of normal RR intervals. SDNN: Standard deviation of normal RR intervals.

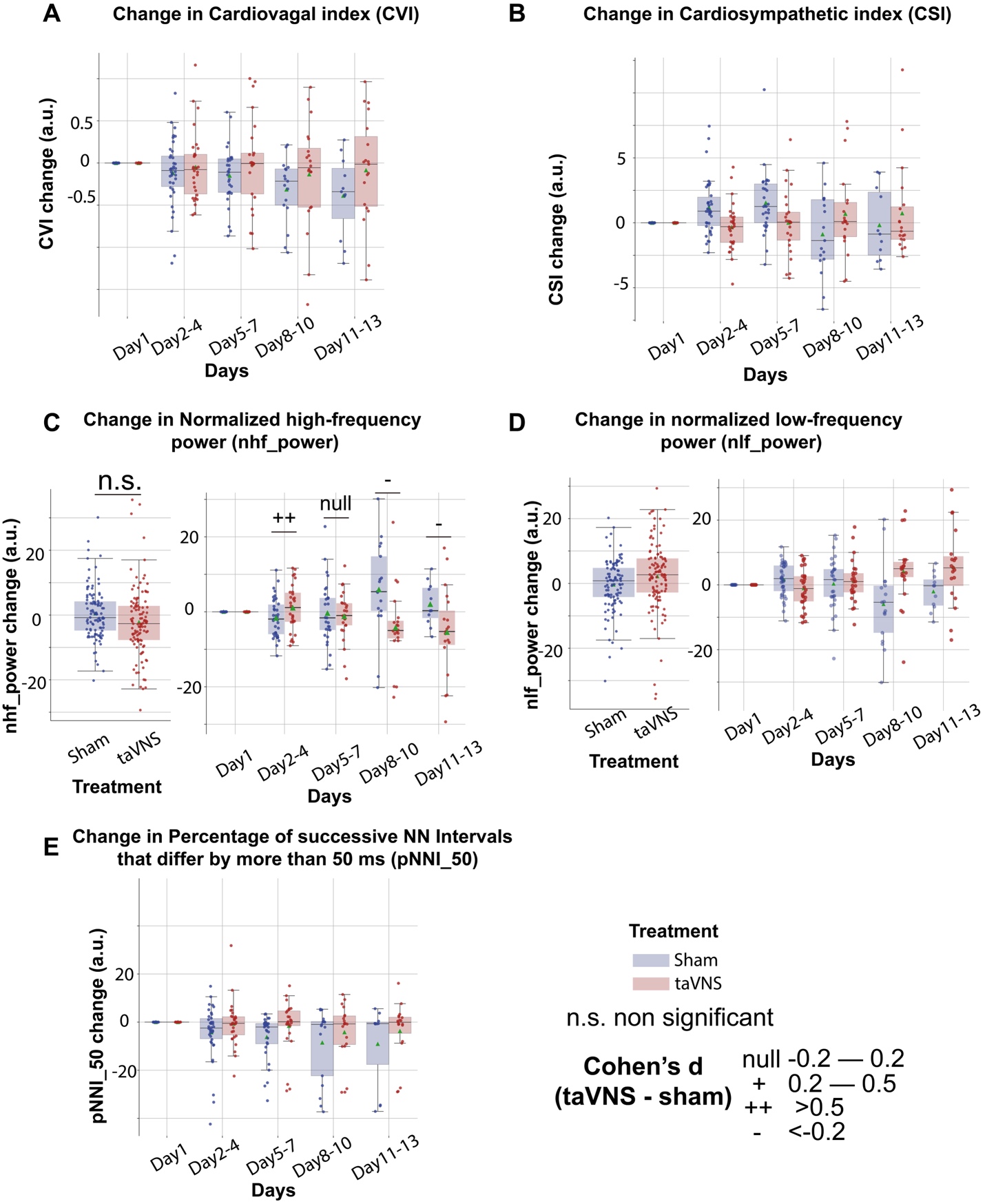

**Supplementary Figure 2** (related to **Figure *3***) **Time-domain and frequency-domain cardiac measures in taVNS and Sham treatment groups.** These standard heart rate variability metrics were used to perform factor analysis.

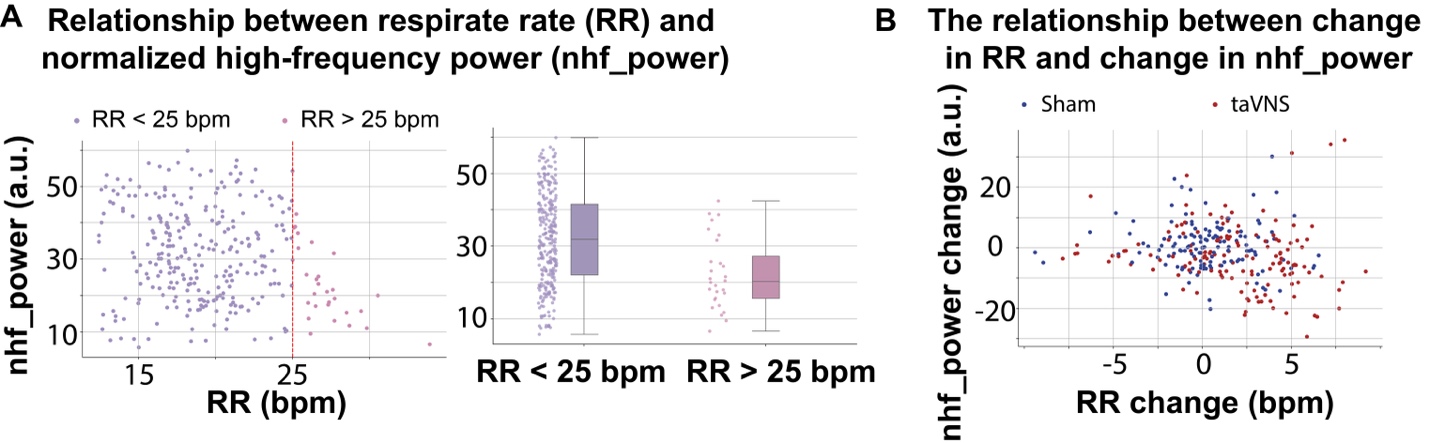

**Supplementary Figure 3** (related to **Figure *3***) **The normalized high-frequency power may not fully represent parasympathetic activity when the respiration rate exceeds 25 bpm. B.** Higher RR change in the taVNS treatment group is associated with less normalized high-frequency power.

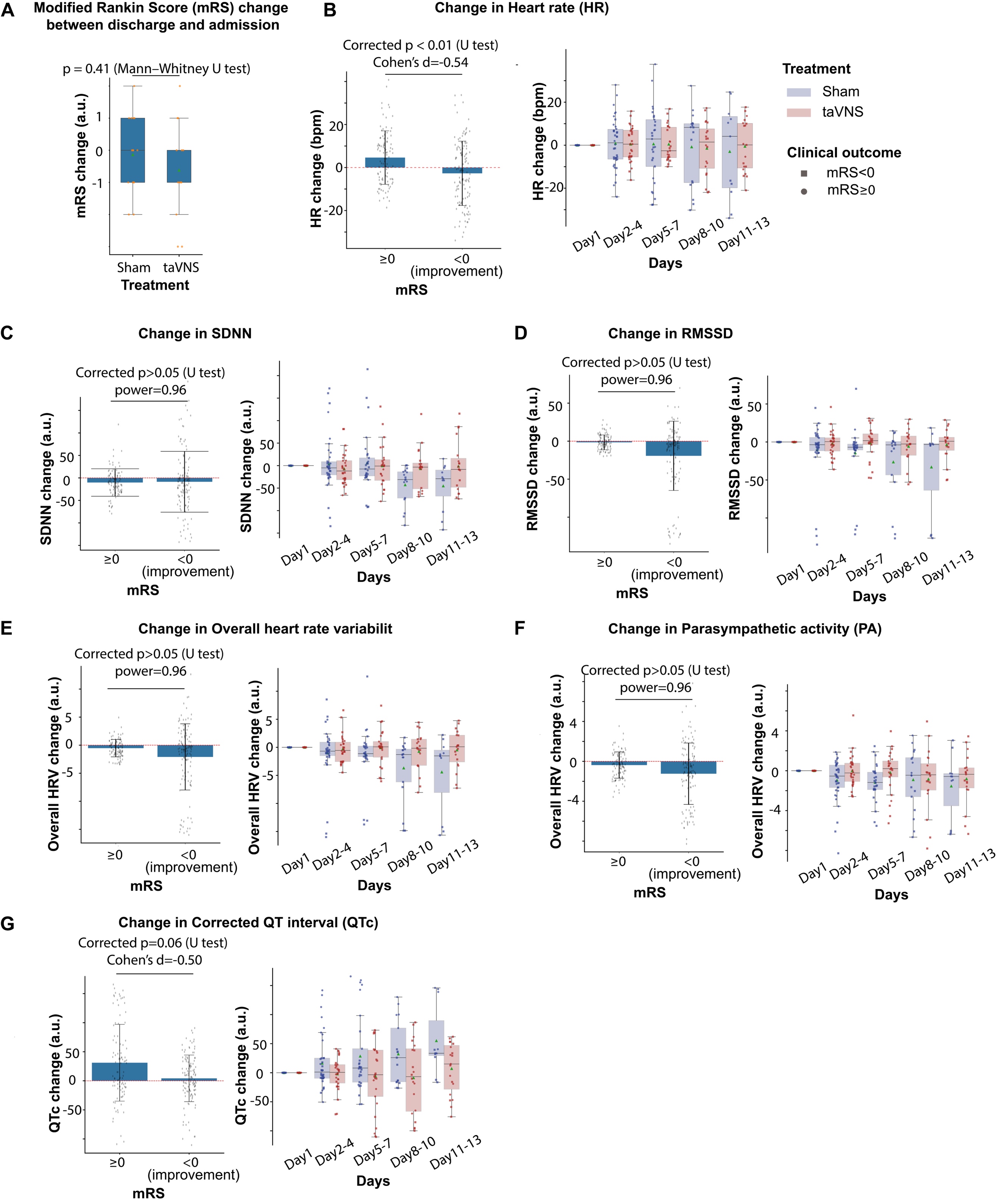

**Supplementary Figure 4** (related to **Figure *3***) **The** **impact of clinical outcome on heart rate variability.** **A**. The change in mRS was similar between the taVNS and Sham treatment groups. **B**. Change in heart rate during treatment for both treatment groups, as compared to first hospitalized day, was lower in patients with improved mRS upon discharge (i.e., mRS change < 0). **C–F**. The relationship between improved mRS and changes in cardiovascular metrics. N(mRS < 0) = 122, N(mRS > 0) = 98.

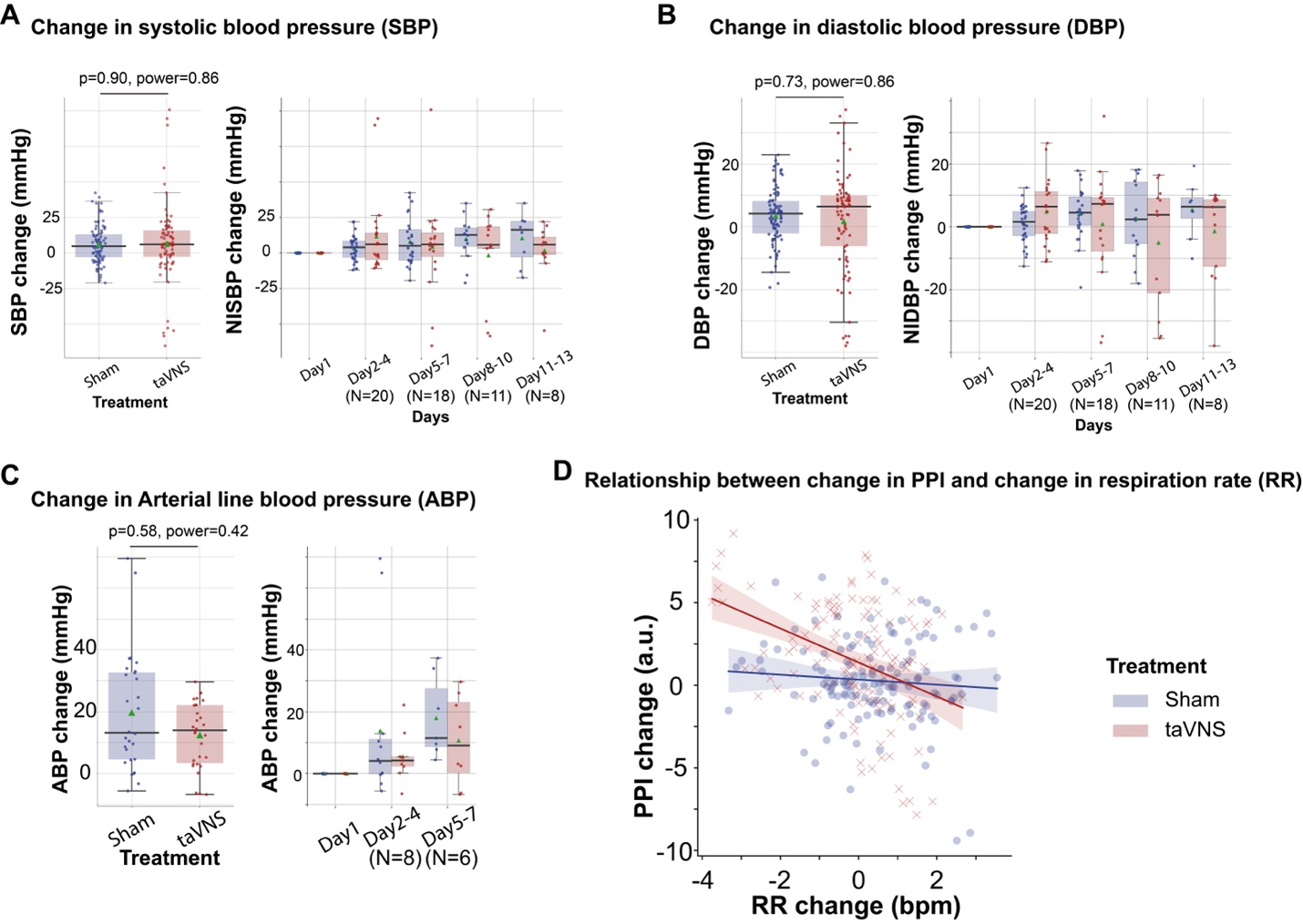

**Supplementary Figure 5** (related to **Figure *4***) **The** **effect of taVNS on arterial line blood pressure monitoring and noninvasive blood pressure monitoring.** Mann–Whitney U tests were used to compare changes in blood pressure between treatment groups.

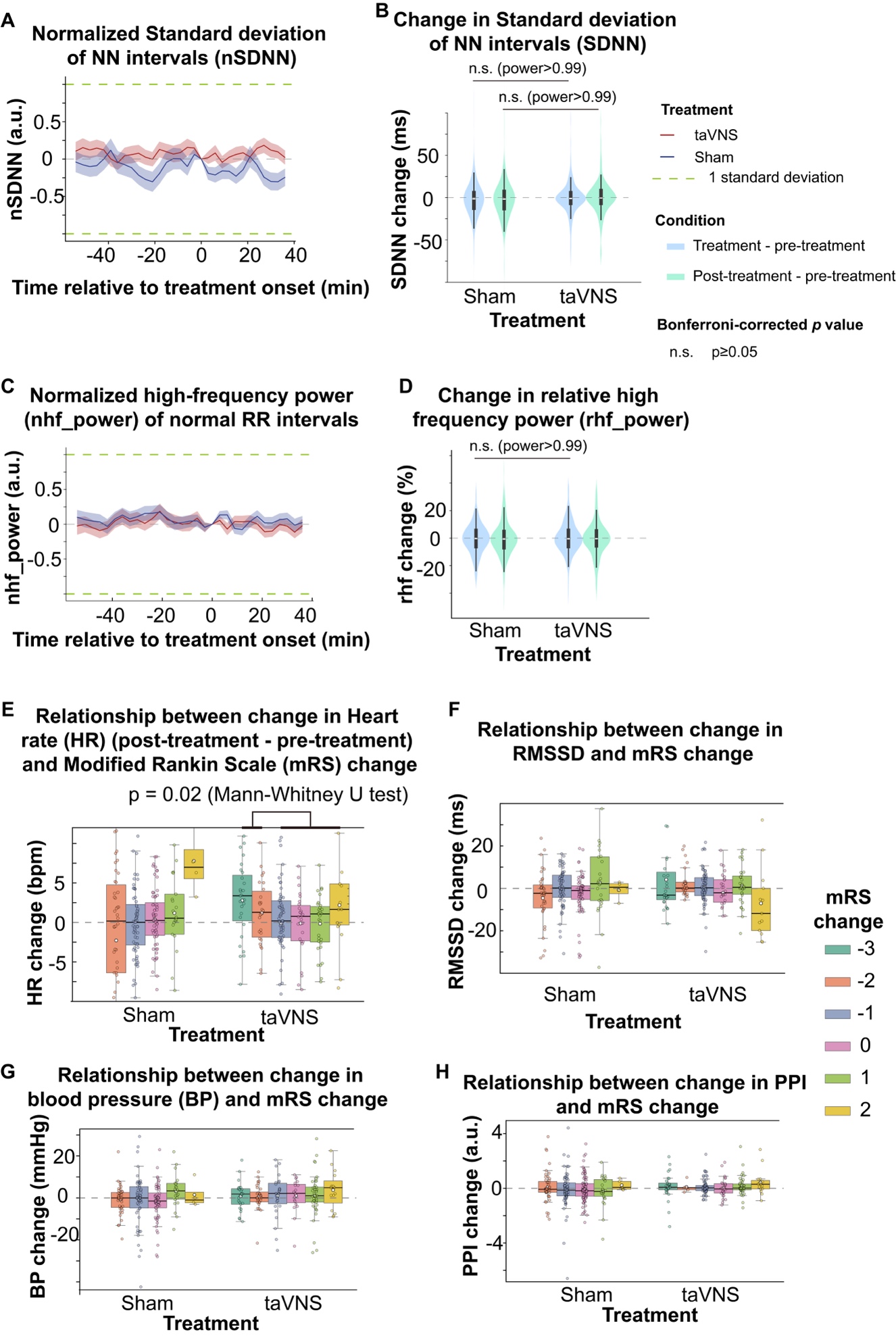

**Supplementary Figure 6** (related to **Figure *5***) **Cardiac effects of acute taVNS.** **A (C)**. Temporal dynamics of normalized SDNN (high-frequency power). Normalized SDNN (high-frequency power) for the two treatment groups at the treatment onset was set to 0. The data is presented as a mean ± standard error. **B (D)**. Comparison of change in SDNN (relative high-frequency power) values from pre-treatment to treatment period (blue) and post-treatment period (green) between treatment groups. No significant differences were observed between treatment groups based on Mann–Whitney U tests. **E-F**. The relationship between changes in the modified Rankin Scale (mRS) scores and changes in differential measures of cardiovascular parameters from pre-treatment to post-treatment periods.

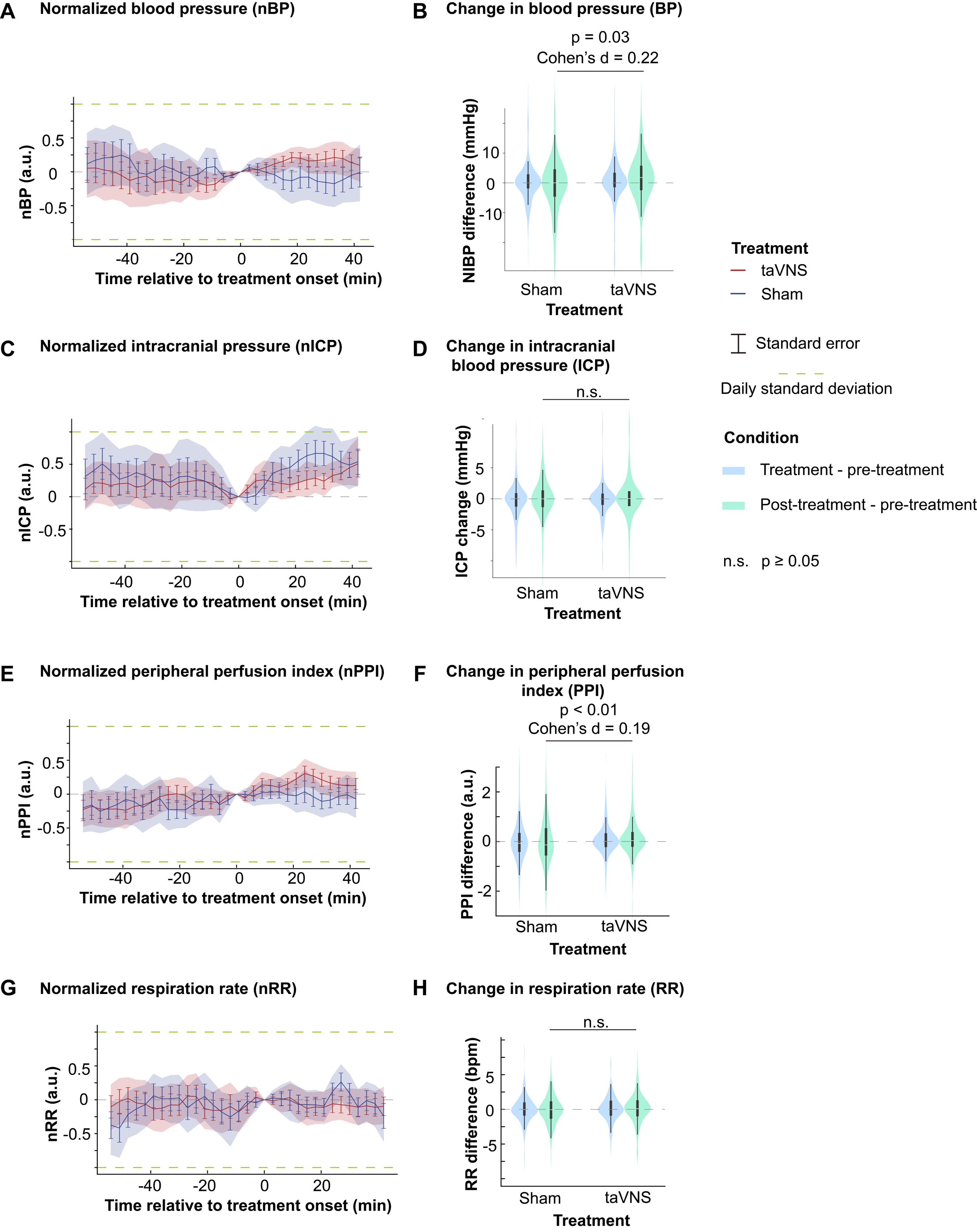

**Supplementary Figure 7** (related to **Figure *5***) **Vascular effects of acute taVNS.** **A, C, E, G (left)**. Temporal dynamics of normalized blood pressure (BP), intracranial blood pressure (ICP), peripheral perfusion index (PPI), and respiration rate (RR) in both treatment groups, aligned with the treatment onset. **B, D, F, H (right).** Comparison of change in BP, ICP, PPI, and RR from pre-treatment to treatment (blue) and post-treatment (green) periods. Mann–Whitney U tests were used to compare the change in vital signs between groups. The powers are > 0.99 for RR and 0.98 for ICP.

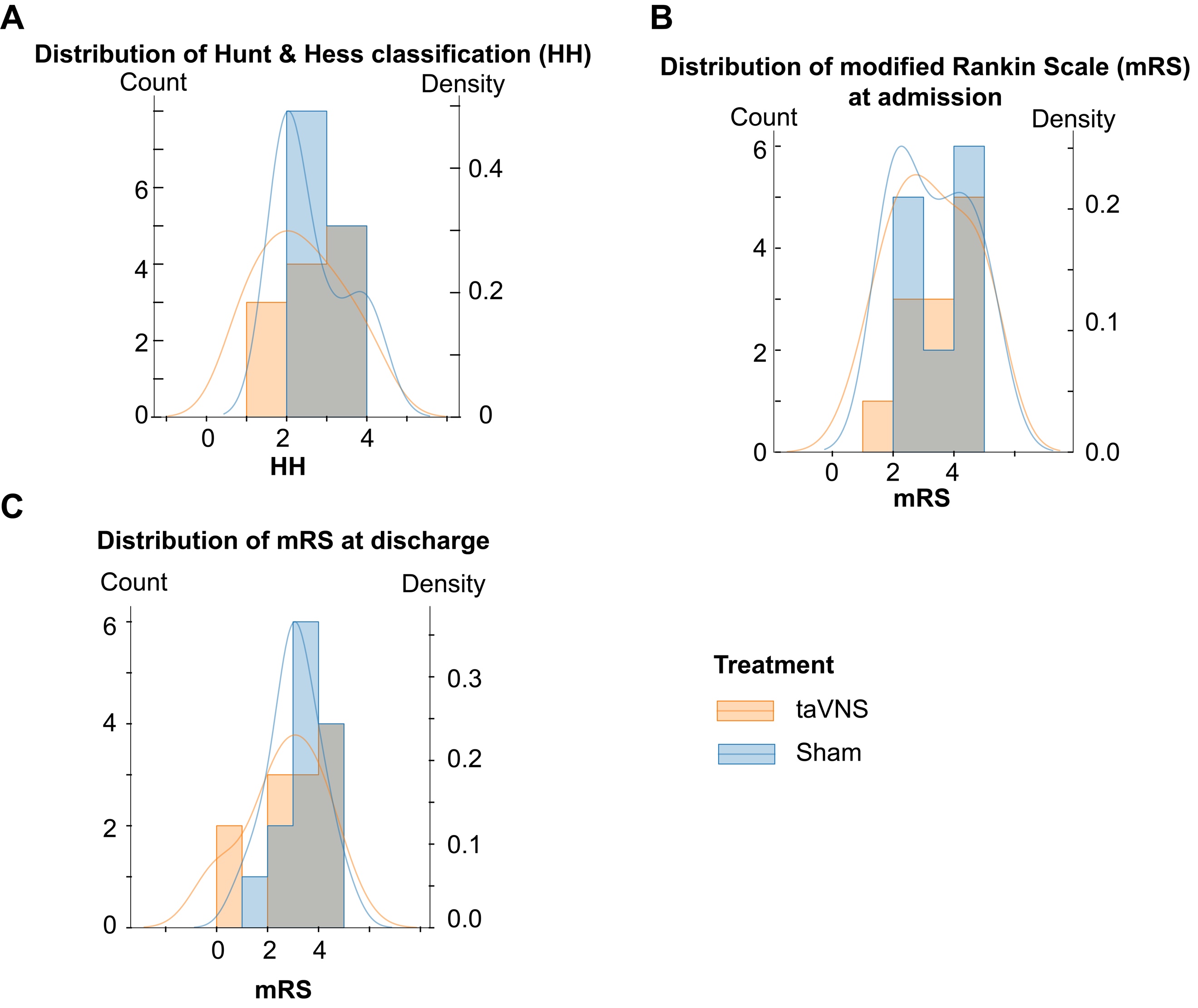

**Supplementary Figure 8** **The distribution of Hunt & Hess classification and modified Rankin Scale for taVNS and Sham groups.**

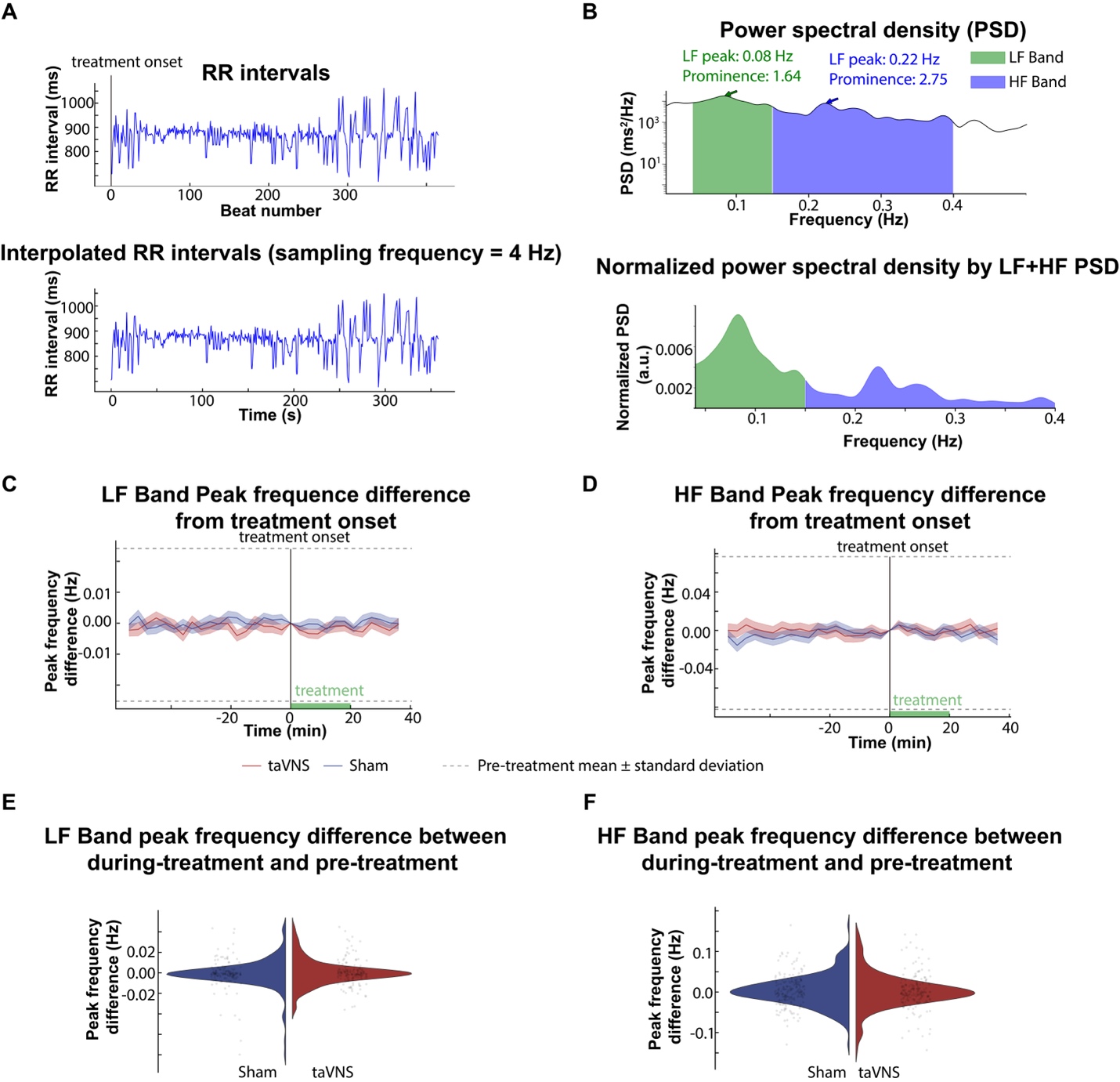

**Supplementary Figure 9** **The effect of acute taVNS on peak frequencies within the high-frequency (HF) band and the low-frequency (LF) band.** To investigate the potential effects of acute taVNS treatment on the autonomic system, we analyzed the peak frequency of the high-frequency and low-frequency bands. **A**. representative RR intervals over 6 minutes were linear interpolated for frequency domain analysis. Ectopic beats and outliers were identified and corrected. **B**. The power spectral density (PSD) of the interpolated RR intervals time series. Peak frequencies within the HF and LF bands and their prominence were calculated from the normalized power spectral density. **C and D**. Changes in LF and HF peak frequencies over time, with peak frequencies at treatment onset (time 0) set as the baseline (0). **E and F**. Comparison of the changes in peak frequencies between the during-treatment and pre-treatment periods across treatment groups (LF band peak frequency: p = 0.54, Cohen's d = -0.07; HF band peak frequency: p-value = 0.67, Cohen's d = 0.04, t-test).

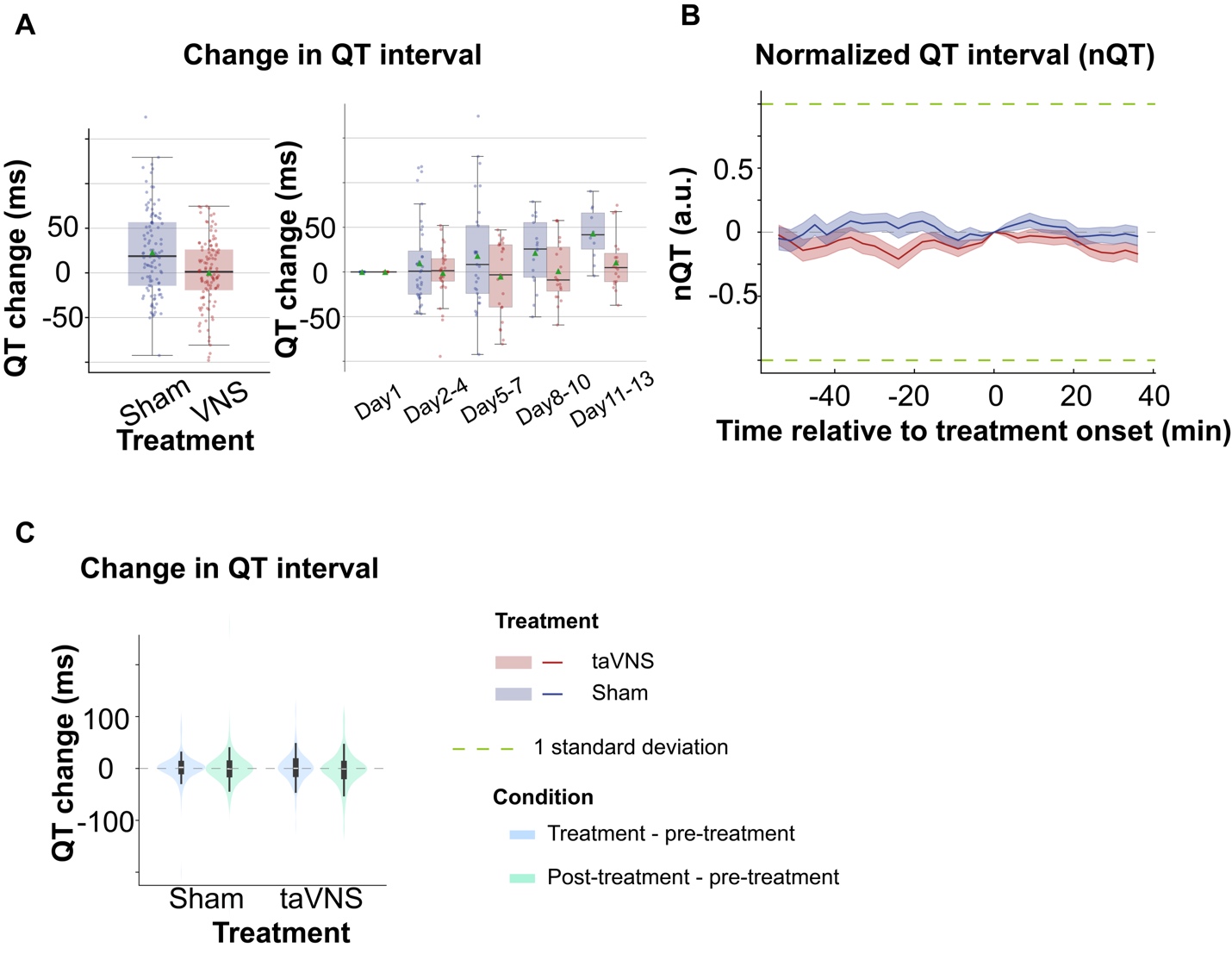

**Supplementary Figure 10** **The effect of repetitive and acute taVNS on uncorrected QT interval. A.** QT interval changes from the first hospitalized day in the two treatment groups. **B**. Normalized QT interval aligned at the treatment onset over time for the two treatment groups. The QT interval is normalized based on the mean and standard error of heart rate for each day. **C**. The difference in QT intervals between the treatment period, post-treatment period, and pre-treatment period for the two treatment groups.

**Supplementary Table 2. Summary of Statistical Tests.** Bolded values were used to make inferences in this paper.

| Test name | Variable | Distribution A | | Distribution B | N(A) | N(B) | p | statistics | Effect size |
| --- | --- | --- | --- | --- | --- | --- | --- | --- | --- |
| T | RMSSD change | taVNS | Sham | | 94 | 95 | **0.004** | t = 2.91 | 0.42 |
| T | SDNN change | taVNS | Sham | | 94 | 95 | 0.48 | t = 0.71 | 0.10 |
| Mann-Whitney U | HR change | taVNS | Sham | | 94 | 95 | 0.69 | U(A) = 4317 | -0.01 |
| Equivalence test (two one sided t-tests) | HR change | taVNS | Sham | | 94 | 95 | **0.006 (lower), 0.004 (upper)** | 2.53 (lower)  -2.72  (upper) | -0.01 |
| Mann-Whitney U | QTc change | taVNS | Sham | | 94 | 95 | **<0.01** | U(A) = 3539 | **-0.57** |
| Equivalence test (two one-sided t-tests) | QTc change  margin:30ms | taVNS | Sham | | 94 | 95 | 0.50 (lower), **1.45***$\boldsymbol{10}^{\boldsymbol{-13}}$ (upper) | -0.004 (lower)  -7.86  (upper) | -0.57 |
| Mann-Whitney U | Percentage of prolonged QT | taVNS | Sham | | 94 | 95 | **<0.01** | U(A) = 2885 | **-0.72** |
| Mann-Whitney U | Overall HRV change | taVNS | Sham | | 94 | 95 | **0.04** | U(A) = 5237 | **0.37** |
| Mann-Whitney U | Parasympathetic activity change | taVNS | Sham | | 94 | 95 | **0.04** | U(A) = 5238 | 0.29 |
| Mann-Whitney U | ICP change | taVNS | Sham | | 66 | 52 | 0.61 | U(A) = 1972 | 0.25 |
| Equivalence test (two one-sided t-tests) | ICP change  margin:2mmHg | taVNS | Sham | | 66 | 52 | **3.66***$\boldsymbol{10}^{\boldsymbol{-13}}$ (lower), **3.33***$\boldsymbol{10}^{\boldsymbol{-10}}$ (upper) | 8.07 (lower)  -6.73  (upper) | 0.12 |
| Mann-Whitney U | BP change | taVNS | Sham | | 66 | 81 | 0.73 | U(A) = 2842 | -0.11 |
| Equivalence test (two one-sided t-tests) | BP change  margin:2mmHg | taVNS | Sham | | 66 | 81 | 0.07(lower), 0.002(upper) | 1.51 (lower)  -3.00  (upper) | -0.12 |
| Mann-Whitney U | SBP change | taVNS | Sham | | 66 | 81 | 0.90 | U(A) = 2719 | -0.07 |
| Mann-Whitney U | DBP change | taVNS | Sham | | 66 | 81 | 0.73 | U(A) = 2846 | -0.17 |
| Mann-Whitney U | ABP change | taVNS | Sham | | 28 | 24 | 0.46 | U(A) = 295 | -0.48 |
| Mann-Whitney U | PPI change | taVNS | Sham | | 83 | 95 | **0.002** | U(A) = 2877 | -0.49 |
| Mann-Whitney U | RR change | taVNS | Sham | | 94 | 95 | **0.004** | U(A) = 5530 | 0.37 |
| Mann-Whitney U | HR difference (post-pre) | taVNS | Sham | | 188 | 199 | 0.28 | U(A) = 17527 | 0.10 |
| Wilcoxon signed rank | HR difference (post-pre) | taVNS |  | | 188 |  | **0.02** | 7525 | 0.11 |
| Mann-Whitney U | QTc difference (post-pre) | taVNS | Sham | | 188 | 198 | 0.86 | U(A) = 18412 | 0.02 |
| Mann-Whitney U | RMSSD difference (post-pre) | taVNS | Sham | | 188 | 199 | 0.31 | U(A) = 17581 | 0.14 |
| Mann-Whitney U | SDNN difference (post-pre) | taVNS | Sham | | 188 | 199 | 0.48 | U(A) = 17923 | 0.03 |
| Mann-Whitney U | HR difference(post-pre) | mRS change<-1 | mRS change$\geq-1$ | | 53 | 122 | **0.01** | U(A) = 4019 | 0.38 |
| Mann-Whitney U | BP difference(post-pre) | taVNS | Sham | | 159 | 180 | **0.03** | U(B) = 12400 | 0.21 |
| Mann-Whitney U | ICP difference(post-pre) | taVNS | Sham | | 146 | 114 | 0.82 | U(B) = 12400 | 0.09 |
| Mann-Whitney U | PPI difference(post-pre) | taVNS | Sham | | 186 | 227 | **0.002** | U(B) = 17386 | 0.19 |
| Mann-Whitney U | RR difference(post-pre) | taVNS | Sham | | 214 | 224 | 0.10 | U(B) = 21806 | 0.11 |

**Supplementary Table 3. Clinical characteristics and cardiovascular metrics of the two arms.** Values in this table represent the median.

|  |  | **taVNS** |  | **Sham** |
| --- | --- | --- | --- | --- |
| Age |  | 67 |  | 53 |
| % of female |  | 63.6% |  | 84.6% |
| % with known hypertension |  | 90.9% |  | 46.2% |
| % with known diabetes mellitus |  | 18.2% |  | 7.7% |
| % with arrhythmia PTA |  | 9.1% |  | 7.7% |
| % with coronary artery disease PTA |  | 0% |  | 15.4% |
| % on beta blockers PTA |  | 27.3% |  | 38.5% |
| % on calcium channel blockers PTA |  | 27.3% |  | 7.7% |
| % on angiotensin-converting enzyme inhibitors PTA |  | 27.3% |  | 15.4 |
| **heart rate (bpm)** |  | 86.0 |  | 78.8 |
| change from pre- to post-treatment |  | 1.1 |  | 0.3 |
| change from pre- to during-treatment |  | 0.4 |  | -0.2 |
| **QTc (ms)** |  | 509.9 |  | 483.6 |
| change from pre- to post-treatment |  | -0.4 |  | -0.2 |
| change from pre- to during-treatment |  | 1.3 |  | 1.0 |
| **SDNN (ms)** |  | 72.7 |  | 77.1 |
| change from pre- to post-treatment |  | -0.4 |  | -1.7 |
| change from pre- to during-treatment |  | -1.0 |  | -1.4 |
| **RMSSD (ms)** |  | 24.8 |  | 24.5 |
| change from pre- to post-treatment |  | -0.1 |  | -0.4 |
| change from pre- to during-treatment |  | -0.3 |  | -0.8 |
| **Relative power of high-frequency band (%)** |  | 30.4 |  | 30.7 |
| change from pre- to post-treatment |  | -0.4 |  | -0.7 |
| change from pre- to during-treatment |  | -0.5 |  | -0.3 |
| **Blood pressure (mmHg)** |  | 92.4 |  | 95.3 |
| change from pre- to post-treatment |  | 1.8 |  | 0.0 |
| change from pre- to during-treatment |  | 0.0 |  | 0.0 |
| **PPI (a.u.)** |  | 1.6 |  | 2.3 |
| change from pre- to post-treatment |  | 0.0 |  | -0.1 |
| change from pre- to during-treatment |  | 0.0 |  | -0.1 |
| **ICP (mmHg)** |  | 6 |  | 6.3 |
| change from pre- to post-treatment |  | 0.0 |  | 0.0 |
| change from pre- to during-treatment |  | 0.0 |  | 0.0 |
| **Respiration rate (bpm)** |  | 20.1 |  | 18.3 |
| change from pre- to post-treatment |  | 0.1 |  | 0.1 |
| change from pre- to during-treatment |  | 0.0 |  | 0.0 |

QTc: Corrected QT interval; PPI: Peripheral Perfusion Index; ICP: Intracranial pressure; PTA: prior to admission.

**Supplementary Table 4. Effect of taVNS on cardiac metrics adjusted for age.** The regression formula is: cardiac metric ~ Treatment + Age.

|  | **Coefficient** | **P value** | **95% CI** |
| --- | --- | --- | --- |
| *Heart rate change from Day 1 (F = 9.49, p < 0.01 for the overall model)* | | | |
| taVNS treatment | 1.87 | 0.21 | [-1.05, 4.80] |
| Age | -0.26 | <0.01 | [-0.37, -0.14] |
| *QTc change from Day 1 (F = 14.23, p < 0.01 for the overall model)* | | | |
| taVNS treatment | -30.79 | <0.01 | [-42.16, -19.43] |
| Age | 0.19 | 0.40 | [-0.26, 0.65] |
| *SDNN change from Day 1 (F = 29.40, p < 0.01 for the overall model)* | | | |
| taVNS treatment | 15.43 | <0.01 | [5.14, 25.72] |
| Age | -1.56 | <0.01 | [-1.97, -1.15] |
| *RMSSD change from Day 1 (F = 25.09, p < 0.01 for the overall model)* | | | |
| taVNS treatment | 16.90 | <0.01 | [10.02, 23.78] |
| Age | -0.83 | <0.01 | [-1.10, -0.56] |
